# Single-cell level characterization of B cell depletion and repopulation following rituximab in systemic lupus erythematosus

**DOI:** 10.1101/2025.05.27.25328230

**Authors:** Haerin Jang, Norzawani Buang, Catherine Sutherland, Wanseon Lee, Lauren Overend, Tarran S. Rupall, Katie L. Burnham, Matthew C. Pickering, Marina Botto, Emma E. Davenport

**Affiliations:** Wellcome Sanger Institute, Wellcome Genome Campus, Hinxton, UK; Department of Immunology and Inflammation, Imperial College London, London, UK; Centre for Human Genetics, University of Oxford, Oxford, UK

## Abstract

**Objective:** Rituximab, a CD20^+^ B cell depletion therapy, is frequently used in the treatment of systemic lupus erythematosus (SLE). However, variability in patient response highlights the need for a deeper understanding of the underlying immune cell dynamics of B cell depletion and repopulation.

**Methods:** In this study, we conducted longitudinal single-cell profiling of nine SLE patients treated with rituximab from pretreatment to up to 15 months post-treatment. These were compared to eight healthy controls. We profiled 169,513 immune cells via single-cell RNA, surface protein, B cell receptor (BCR), and T cell receptor (TCR) sequencing and sequenced bulk BCR repertoires in parallel.

**Results:** Significant depletion of naïve, memory, and age-associated B cells (ABCs) was observed early post-treatment, followed by later repopulation of mainly transitional B cells. BCR repertoire analysis revealed reduced diversity and persistent clones in antigen-experienced cells at early post-treatment, but these effects were not long-lasting. Notably, repopulated naïve B cells in rituximab responders exhibited reduced NF-κB pathway activation, aligning with lower BAFF-R surface protein levels. In non-B cells, we identified 27 differentially expressed genes across 7 immune cell subtypes post-rituximab, with regulatory CD4 T cells and double negative (DN) T cells showing the most transcriptional changes. Responders specifically had increased expression of genes related to cytotoxicity, MHC class II antigen presentation, and T cell activation in CD4 T central memory (TCM) and DN T cells.

**Conclusion:** Our longitudinal profiling provides single-cell resolution of the shifts in immune cell dynamics following B cell depletion.

## Introduction

Systemic lupus erythematosus (SLE) is a chronic autoimmune disease characterized by widespread inflammation and tissue damage in multiple organs, including the skin, kidneys, joints, and central nervous system. SLE is driven by dysregulated immune responses, notably aberrant B cell activity promoting the production of autoantibodies targeting self-antigens^1^. Historically, treatment relied on broad immunosuppressants, which often carried significant side effects. However, advances in our understanding of SLE pathophysiology have led to the development of targeted biologic therapies such as anti-BAFF, anti-CD20, and anti-IFN-α receptor antibodies^2^. By focusing on specific molecular pathways and cell types involved in disease progression, these therapies offer more targeted and potentially safer treatment options, while reducing reliance on broad immunosuppressants. However, our incomplete understanding of treatment mechanisms has led to limited success in clinical trials.

Rituximab, an anti-CD20 monoclonal antibody, is frequently prescribed off-label for SLE^3^ despite failing to meet endpoints in clinical trials^4,5^. It depletes CD20^+^ B cells in peripheral blood, followed by gradual repopulation over months. By removing B cells, rituximab likely reduces autoantibody production and release of proinflammatory cytokines^6^, though its precise mechanisms in achieving symptom relief are incompletely understood. Similarly, it is unclear what underpins the variability in patient response that is observed^3^. With emerging B cell depletion therapies and the recent success of obinutuzumab, a newer anti-CD20 monoclonal antibody, in lupus nephritis^7^, understanding changes in the immune response following these treatments is increasingly important.

Previous studies have examined specific B cell subpopulations following rituximab treatment using flow cytometry^8,9^ and bulk transcriptomics^10^, but molecular changes within peripheral blood mononuclear cells (PBMCs) using single-cell technologies remain unexplored. Unlike bulk RNA-seq methods, single-cell RNA-seq (scRNA-seq) reveals cell-type-specific transcriptome changes as well as shifts in cell abundance. This approach is particularly advantageous for studying immune cells, where gene function is highly cell-type-specific. Studying B cell depletion therapy with scRNA-seq offers insights into the dynamic changes in not only the B cell subtypes targeted but also non-B cells, helping to better understand the broader immune response and potential off-target effects. For example, a single-cell study on CD19 CAR-T cell mediated B cell depletion in SLE found reduced type I interferon-induced gene expression in monocytes and T cells, but not in B cells^11^.

Here, we characterized B cell depletion and repopulation following rituximab in SLE patients at the single-cell level. Using a multimodal single-cell approach comprising gene expression (scRNA-seq), surface protein level measurements (CITE-Seq: cellular indexing of transcriptomes and epitopes) and B cell receptor (BCR) and T cell receptor (TCR) repertoire data, we analyzed changes in immune cells in nine SLE patients. Combined with bulk BCR data, we observed dynamic shifts in B cell subtypes after depletion and repopulation, while the BCR repertoire remained largely unchanged long-term. By further stratifying the patients into responders and non-responders to rituximab, we found that repopulated naïve B cells in responders are transcriptionally distinct from pretreatment and observed an increase in cytotoxicity and activation related genes in responder central memory CD4 T cells (CD4 TCM) and double negative (DN) T cells. Our findings offer a detailed view of immune cell dynamics in SLE patients receiving rituximab.

## Materials and Methods

### Sample collection and sequencing

Blood samples from SLE patients receiving rituximab were collected from patients ≥18 years old at the Imperial Lupus Centre, Imperial College Healthcare NHS Trust, UK. None of the SLE patients had received rituximab in the 6 months before enrolment. ‘Early post-treatment’ samples were collected 1 to 6 months after treatment and ‘Late post-treatment’ samples 7 to 15 months after treatment. Response was determined by a clinician at 12 months post-treatment following the criteria outlined in **Supplementary Methods**. Blood samples from healthy controls were also collected. Participant ancestry was self-reported from a fixed set of categories. Human samples used in this research project were obtained from the Imperial College Healthcare Tissue and Biobank (ICHTB). ICHTB is supported by the National Institute for Health Research Biomedical Research Centre based at Imperial College Healthcare NHS Trust and Imperial College London. ICHTB is approved by Wales REC3 to release human material for research (22/WA/0214) and the samples for this project (Ref: R13010-3A) were issued from subcollection reference number IMM_MB_15_027 and IMM_MB_13_001. Written informed consent was obtained from all participants.

From collected blood samples, PBMCs were isolated and cryopreserved. Cells were processed using the 10x Chromium platform to generate gene expression, surface protein (n=137), TCR and BCR libraries, then sequenced using NovaSeq 6000 (Illumina). In addition, bulk BCR library preparation and sequencing followed the protocol from Bashford-Rogers et al.^12^ Details on library preparation and sequencing are in **Supplementary Methods**.

### Data preprocessing

A total of 256,696 cell-containing droplets were obtained using Cell Ranger multi v7.0.0 (10x Genomics). QC steps were taken to remove low-quality cells, leaving 169,513 cells (**Supplementary Methods, Supplementary Fig. 1**). Filtered BCR and TCR contigs from Cell Ranger were processed using the Immcantation framework^13^ and IgBLAST^14^ (v1.21.0). 13,918 BCR and 67,062 TCR sequences were used in downstream repertoire analyses.

Multimodal integration between scRNA-seq and CITE-seq data was performed with totalVI^15^. The resulting low-dimensional latent representations were used for neighborhood construction (k=15) and uniform manifold approximation and projection (UMAP) generation. Unsupervised clustering was performed using the Leiden algorithm^16^. Cell type annotation was determined by canonical RNA and surface protein markers, with B cell annotation additionally informed by isotype usage and mutation frequency (**Supplementary Fig. 2,3**). Normalized CITE-seq values from totalVI were used for analyses on surface protein levels. Details on single-cell data preprocessing are in **Supplementary Methods**. Raw bulk BCR sequence data were processed using the Immcantation framework and IgBLAST v1.21.0 (**Supplementary Methods**). Due to technical issues, IGHV4 sequences were either not amplified or were removed during post-processing. In total, 1,028,618 unique sequences passed QC.

### Differential abundance analysis

Differentially abundant cell populations between timepoints were identified using MiloR (v.2.0)^17^. We compared the cell enrichment at each timepoint within each cell neighborhood using a mixed effect model adjusting for individual patient effects. For B cell subtype-specific changes, neighborhoods were constructed on B cells alone. Spatial FDR < 0.1 was used to determine significance. Analysis methods are detailed in **Supplementary Methods**.

### BCR and TCR repertoire analysis

OLGA^18^ was used to calculate the generation probability of nucleotide CDR3 sequences. Repertoire diversity was assessed by calculating Shannon Entropy on size-matched samples using the *dit* Python package^19^ (**Supplementary Methods**). Heavy chain single-cell and bulk BCR data from all samples for each patient were combined to identify clonal lineages across timepoints. Clustering of sequences into clonal groups was carried out using Change-O^20^. Clones were considered persistent if they contained sequences from both pretreatment and one post-treatment timepoint. Clones containing only sequences from pretreatment were considered non-persistent. Throughout, to account for varying numbers of antibody secreting cells, bulk BCR sequences were not weighted by UMI counts.

### Differential gene expression analysis and pathway analysis

We used a pseudobulk approach, aggregating raw counts by sample and cell type, to identify differentially expressed genes (DEGs). Trimmed mean of M values (TMM) normalization was performed on highly expressed genes and a quasi-likelihood approach from edgeR^21^ was used. As covariates, the model included patient and cellular detection rate (the fraction of detected genes per cell to adjust for sequencing depth as described in Soneson et al^22^). FDR < 0.05 was used to determine significance.

We tested only cell types with >10 cells in all of the samples compared to ensure that the pseudobulk expression accurately represented the gene expression profile of the cell type. For non-B cells, we first compared pretreatment to early post-treatment in all patients. To further investigate changes in interferon-stimulated genes in these cells, we scored cells using ‘scanpy.tl.score_genes’ from Scanpy (v1.10.1^23^) with ‘Interferon signaling’ pathway genes from Reactome (R-HSA-913531).

To identify differences in depleted and repopulated B cells, we conducted a differential gene expression analysis in naïve B cells from responders at pretreatment and late post-treatment. Gene set enrichment analysis was performed with Reactome pathways (ReactomePA v1.48.0, reactome.db_1.88.0^24^) using the resulting log fold change (FC)-ranked list of genes (including both up- and down-regulated genes). From the most significant results, we assessed the direction and magnitude of NF-κB pathway activation in naïve B cells using the PROGENy database^25^. Pathway activation scores were calculated per cell using decoupleR (v.2.8.0)^26^.

To identify genes in non-B cells where response status altered expression changes, we implemented the approach detailed in the edgeR User’s Guide (Section 3.5). This identifies genes where changes in expression between timepoints differ depending on response. To confirm these results were robust, a permutation analysis was conducted. Detailed methods are available in **Supplementary Methods**.

## Results

### Longitudinal single-cell profiling of rituximab response in SLE

We conducted longitudinal single-cell profiling in 9 SLE patients treated with rituximab to characterize immune cell dynamics following B cell depletion (**Fig. 1a**). PBMC samples were collected at pretreatment and multiple timepoints up to 15 months post-treatment to capture initial B cell depletion and later repopulation. Samples were also collected from 8 healthy controls to provide a reference. Participant demographics are outlined in **Supplementary Table 1 and 2**.

**Figure 1.**
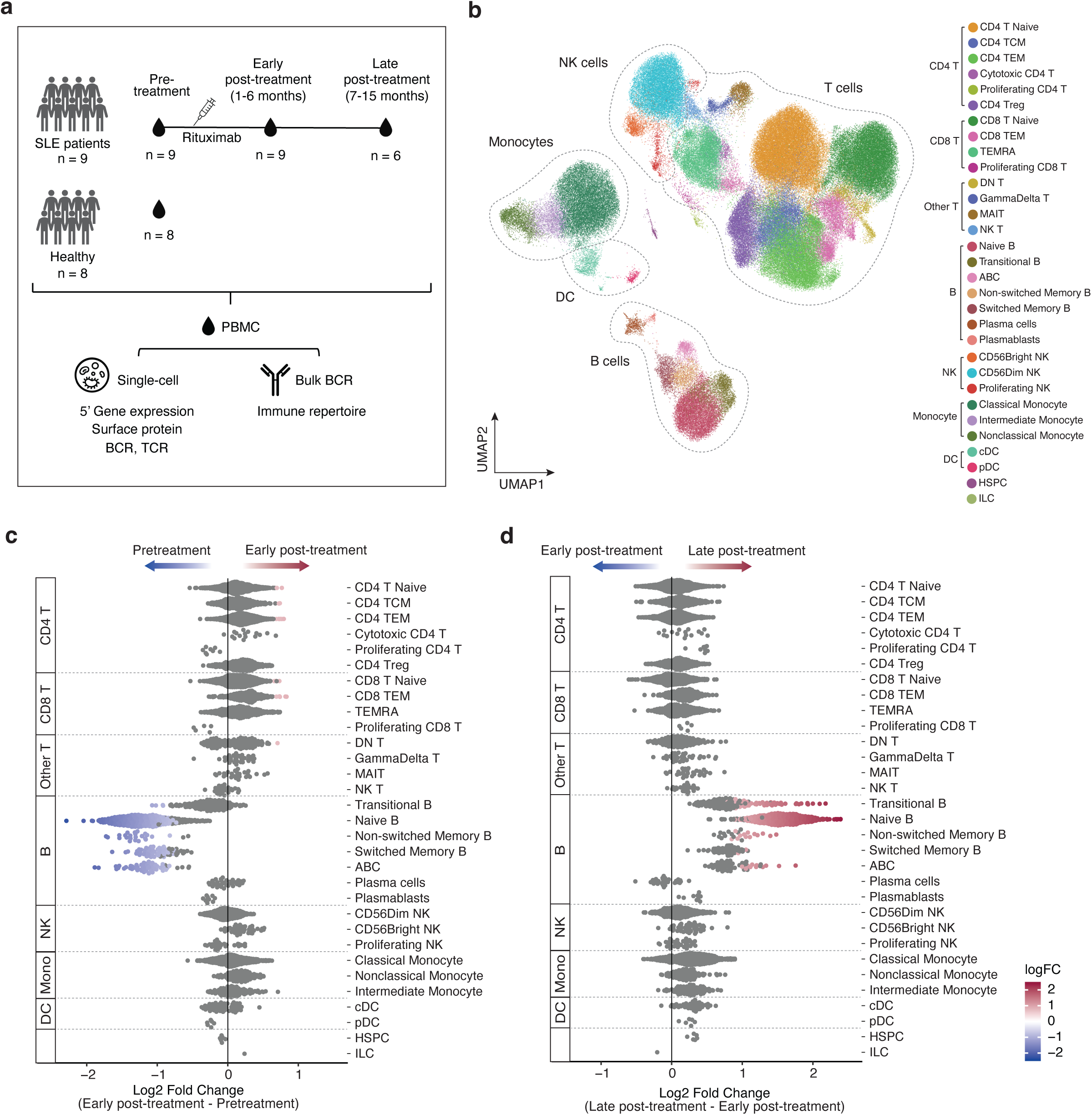
Longitudinal single-cell profiling of response to rituximab in SLE. **a)** Overview of study design. PBMCs were collected from 9 SLE patients and 8 healthy controls at three time points: pretreatment, early post-treatment, and late post-treatment. scRNA-seq, CITE-seq (surface protein profiling), single-cell BCR/TCR sequencing and bulk BCR sequencing were performed. Pretreatment bulk BCR repertoire data were available for 8 out of 9 SLE patients. **b)** UMAP of 169,513 cells, annotated into 31 immune cell types. **c, d)** Differential cell abundance between (c) pretreatment and early post-treatment (n=9 vs 9) and (d) early post-treatment and late post-treatment (n=9 vs 6). Each point represents a cell neighborhood, where most cells belong to the annotated subtype. Neighborhoods with significantly different cell abundances between time points are colored according to their log_2_FC.

A total of 169,513 immune cells were profiled with single-cell RNA, surface protein, BCR, and TCR sequencing. After clustering, we annotated 31 immune cell subtypes using canonical markers (**Fig. 1b, Supplementary Fig. 2)**. B cell subtype annotation was additionally informed by BCR isotype usage and mutation frequency (**Supplementary Fig. 3**).

To assess B cell depletion and repopulation after rituximab, we conducted differential cell abundance analysis between early post-treatment and pretreatment (n=9 vs 9, **Fig. 1c**). Naïve B cells, non-switched memory B cells, switched memory B cells and age-associated B cells (ABCs defined as CD11c^+^, Tbet^+^, CD21^low^, CD27^low^) were significantly less abundant at early post-treatment. Other cell types remained largely unchanged. This included transitional B cells, suggesting rapid repopulation of this cell type given the rapid turnover and limited lifespan of this cell type^27^. Minor increases in CD4 and CD8 T cell neighborhoods are likely due to relative increase in cell capture due to the extensive B cell depletion, rather than an actual expansion. This increase was not confined to a specific cell subtype and was observed across all patients. Comparing late post-treatment to early post-treatment, transitional B cells, naïve B cells, non-switched memory B cells and ABCs increased, reflecting full repopulation (n=9 vs 6, **Fig. 1d)**.

### Perturbation of the BCR repertoire after rituximab

We performed both single-cell V(D)J enrichment and bulk heavy chain BCR sequencing to profile the immune receptor repertoire pre- and post-rituximab. 13,981 paired BCR and 1,028,618 unique heavy chain sequences were identified. To explore repertoire characteristics, we calculated Shannon Entropy, a measure of diversity; generation probability, the likelihood of a sequence being generated by random chance; and somatic hypermutation frequency (**Fig. 2a,b,c**). At pretreatment, these metrics were similar across patients. At early post-treatment, diversity and generation probability decreased in most patients, whilst mutation frequency increased. Patients with poor B cell depletion generally maintained a more diverse repertoire (**Supplementary Fig. 4a**). By late post-treatment, these metrics returned towards pretreatment levels. Isotype usage also varied across timepoints, with increased class-switched sequences at early post-treatment and a reduction at late post-treatment. This is consistent with the reduction and subsequent repopulation of naïve B cells (**Fig. 2d**).

**Figure 2.**
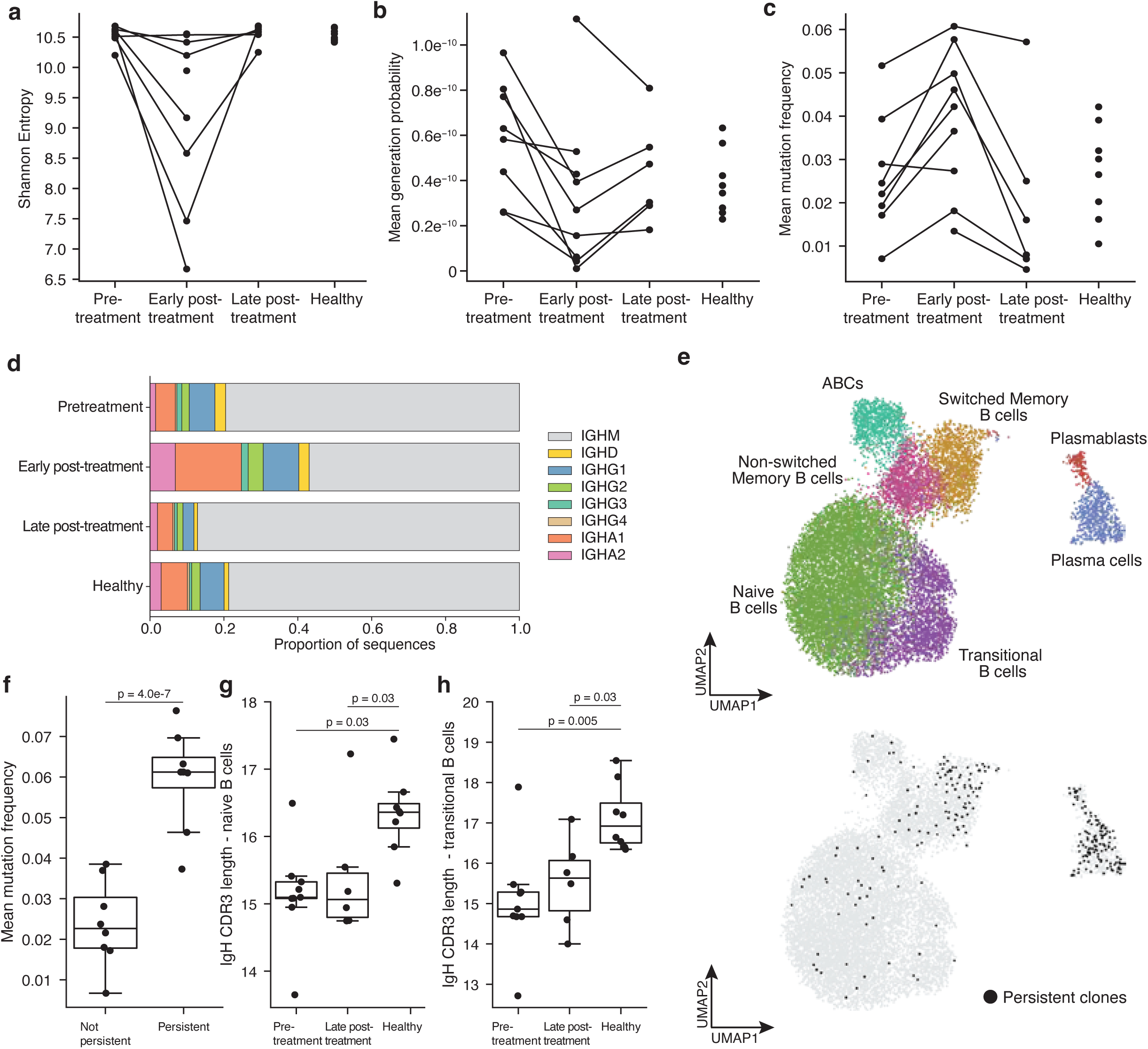
Perturbation of the BCR repertoire after rituximab. **a)** Shannon entropy of bulk BCR repertoire samples across timepoints. Higher values indicate greater diversity. All samples were subset to 1660 UMIs across 1000 iterations and a mean was calculated. **b)** Mean generation probability of bulk BCR repertoire samples across timepoints, calculated by OLGA on nucleotide CDR3 sequence. Higher generation probability indicates a sequence that is more likely to occur by chance. **c)** Mean mutation frequency of bulk BCR repertoire samples across timepoints. **d)** Isotype usage in total B cells at each timepoint from single cell data. **e)** B cell UMAP highlighting cells with sequences belonging to persistent clones. Clones were considered persistent if they contained sequences from both pretreatment and one post-treatment timepoint. **f)** Mean mutation frequency of sequences belonging to persistent or non-persistent clones. Paired t-test. **g,h)** Mean IgH CDR3 length at pretreatment, late post-treatment and in healthy controls in (g) transitional B cells and (h) naïve B cells. FDR adjusted p-value, Kruskal-Wallis followed by Dunn’s test.

BCR clones spanning multiple timepoints were observed in all patients, although the degree of overlap varied (**Supplementary Fig. 4b**). These persistent clones were enriched for switched memory B cells, plasmablasts, and plasma cells (**Fig. 2e, Supplementary Fig. 4c**). Persistent clones had higher mean mutation frequency (paired t-test, p = 4.0e-7, **Fig 2f**). Together, this suggests depletion of less mature B cells and persistence of antigen experienced cells.

As transitional and naïve B cells are preferentially depleted and subsequently repopulated, we examined the BCRs expressed by these cells. Interestingly, SLE patients had shorter CDR3 lengths in both transitional and naïve B cells compared to controls, and this was consistent at pretreatment and late post-treatment (Kruskal-Wallis with Dunn’s test, adjusted p = 0.03 for both, **Fig. 2g,h**). SLE patients also showed biased IGHJ gene usage, particularly in IGHJ3 and IGHJ6 in naïve B cells (**Supplementary Fig. 4d**). Similar signatures were observed in IgM^+^/IgD^+^ SHM^-^ sequences from bulk data (**Supplementary Fig. 4e,f**).

Single-cell TCR data showed no consistent differences in repertoire characteristics after rituximab (**Supplementary Fig. 5**).

### Transcriptomic changes in non-B cells after rituximab

Given that rituximab may lead to change in other immune cells, we analyzed transcriptional changes following rituximab in non-B cells. We performed differential gene expression analysis in non-B cell subtypes with >10 cells in all patients at each timepoint and compared pretreatment to early post-treatment cells to identify early changes after rituximab. A total of 27 genes (26 unique) were identified across 7 out of 10 immune cell subtypes (**Fig. 3a,b**). Regulatory CD4 T (Treg) cells and DN T cells had the most DEGs with 9 and 8 genes, respectively.

**Figure 3.**
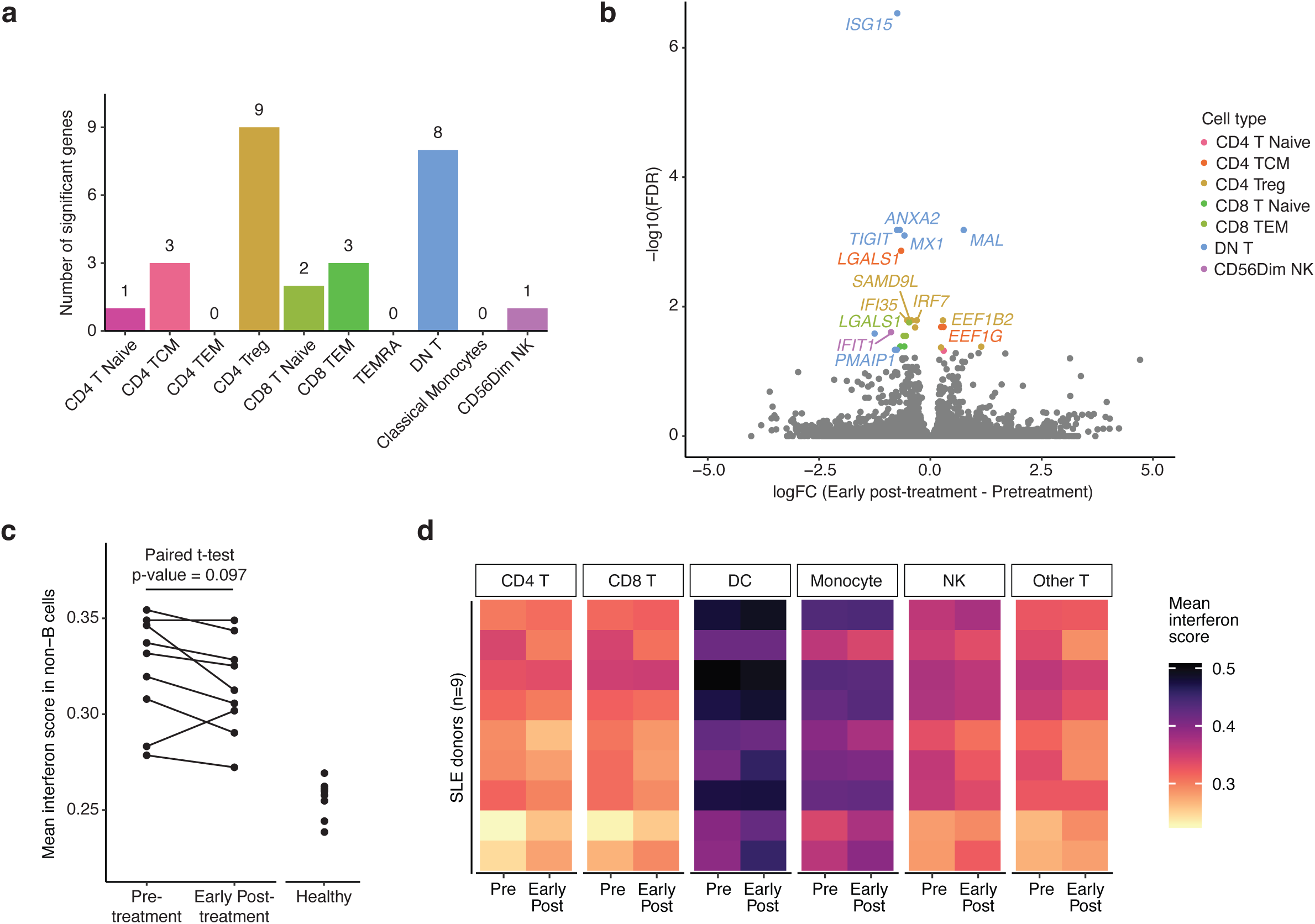
Gene expression changes in non-B cells following rituximab. **a)** Number of significant DEGs in non-B cells between pretreatment and early post-treatment (FDR < 0.05, n=9 vs. 9). Only cell types with at least 10 cells in all patients at both time points were included in the analysis. **b)** DEGs between early post-treatment and pretreatment. Statistically significant results are colored by cell type. **c**) Mean interferon score of non-B cells at pretreatment and early post-treatment to rituximab. Interferon score was calculated using genes from the Reactome gene set ‘Interferon signaling (R-HSA-913531)’. Healthy control scores are shown as reference. **d)** Mean interferon score of each non-B cell subtype at pretreatment and early post-treatment by patient.

Given the known role of interferon signaling in driving inflammation, autoantibody production, and disease prognosis in SLE^28^, we focused on transcriptomic changes in interferon-related genes. Out of the 26 DEGs, eight genes were within the ‘interferon signaling’ pathway (Reactome pathway R-HSA-913531): IFI35, XAF1, and IRF7 in Treg cells, IFI44 in effector memory CD8 T cells, USP18 in naïve CD8 T cells, IFIT1 in CD56Dim NK cells, and ISG15 and MX1 in DN T cells. For most cases, expression decreased following treatment (**Supplementary Fig. 6a**). To assess overall pathway activity, we calculated a score for each cell using ‘interferon signaling pathway’ genes from Reactome (R-HSA-913531) (**Methods**). No consistent changes in interferon pathway scores were observed in non-B cells globally or at the cell-type level (**Fig. 3c,d, Supplementary Fig 6b**).

### B cell depletion and repopulation in rituximab responders and non-responders

To assess potential single-cell-level differences based on the clinical response, we examined cell-type abundance over time in rituximab responders and non-responders separately (**Fig. 4a**). Response was assessed 12 months after treatment (**Supplementary Methods**), with 5 out of 9 SLE patients responding to rituximab.

**Figure 4.**
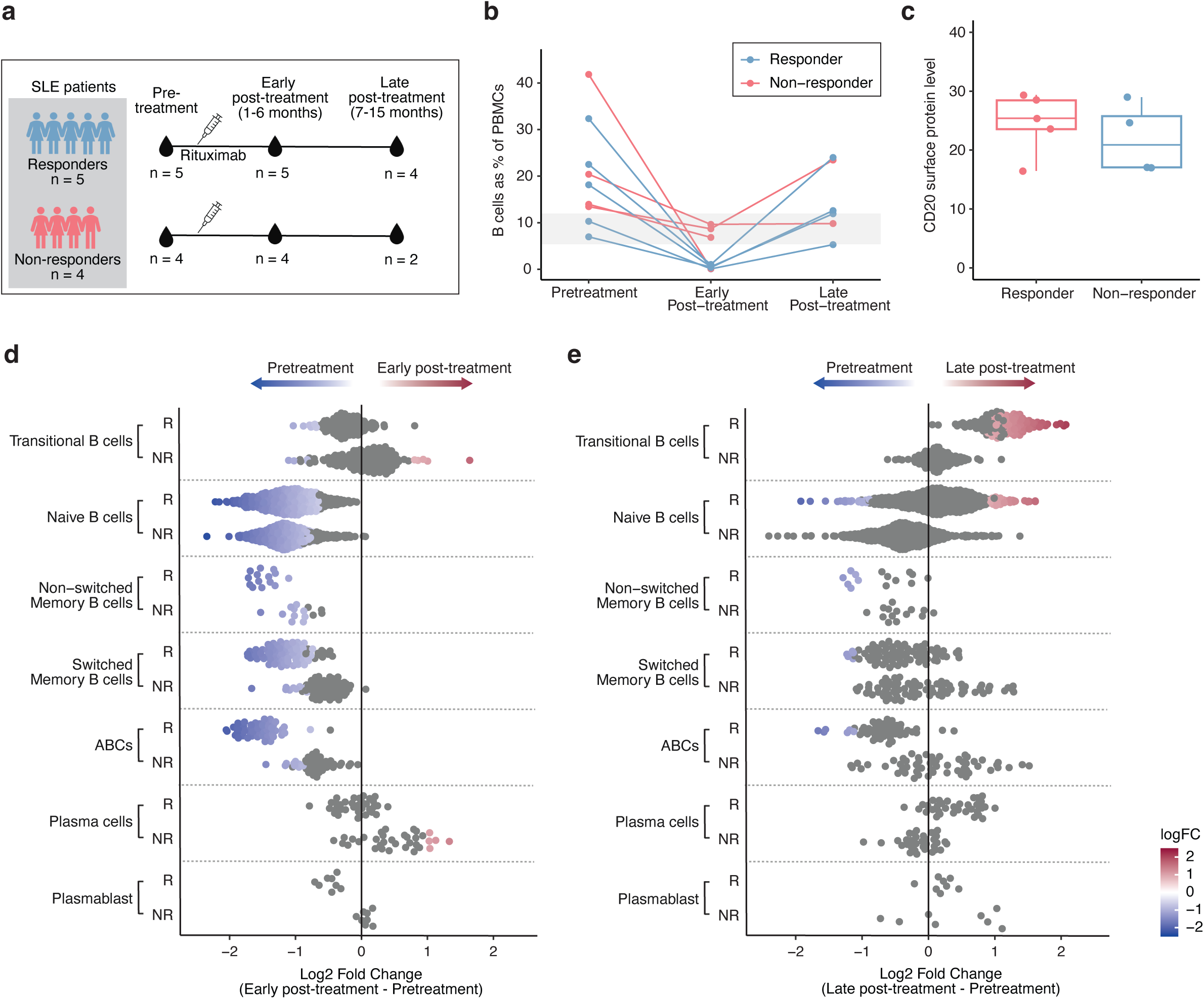
B cell depletion and repopulation in rituximab responders and non-responders. **a)** SLE patients stratified into responders (n=5) and non-responders (n=4) based on clinical response to rituximab. Early post-treatment samples were collected for all patients, while late post-treatment samples were available for 4 out of 5 responders and 2 out of 4 non-responders. **b)** Proportion of B cells among total PBMCs at pretreatment, early post-treatment, and late post-treatment, as measured by single-cell sequencing. The shaded region represents the range observed in healthy controls (n=8). **c)** Average CD20 surface protein expression in B cells from rituximab responders and non-responders at pretreatment. **d,e)** Differential B cell abundance in rituximab responders (R) and non-responders (NR). (d) Comparison between pretreatment and early post-treatment; (e) comparison between pretreatment and late post-treatment. Each point represents a cell neighborhood predominantly composed of the annotated B cell subtype. Neighborhoods with significant changes in abundance are colored by the log_2_FC between timepoints.

To quantify overall B cell depletion, we first calculated the proportion of B cells in single-cell PBMC data pre- and post-treatment (**Fig. 4b**). The B cell proportion was higher than the range found in healthy controls for 7/9 SLE patients at pretreatment, but no difference existed between responders and non-responders (Wilcoxon test p-value > 0.05). All patients showed reduced B cell proportions after treatment, but incomplete depletion (>1% B cells at early post-treatment) was observed in 3/4 non-responders, with an average of 8.4% of PBMCs remaining as B cells. This incomplete depletion was not due to differences in CD20 surface protein levels at pretreatment (t-test p-value > 0.05, **Fig. 4c, Supplementary Fig. 7**). By late post-treatment, B cell proportions were not significantly different from pretreatment (Repeated measures ANOVA p-value > 0.05, **Fig. 4b**).

To determine the difference between responders and non-responders in B cell depletion, we performed differential cell abundance analysis between pretreatment and early-post treatment within each group (**Fig. 4d**). This revealed significant depletion in ABCs and switched memory B cells in responders that was not found in non-responders, suggesting a difference in the effectiveness of depletion in these cells. Naïve B cells and non-switched memory B cells depleted similarly, while non-responders showed a slight increase in plasma cells.

We then compared late post-treatment to pretreatment to understand B cell repopulation (**Fig. 4e**). Responders maintained low ABC and switched memory B cell levels, while transitional B cells rapidly repopulated. Consistent with these changes, BCR data showed IgM usage predominating at late post-treatment (**Supplementary Fig. 8a**). In non-responders, no significant differences in abundance were observed between pretreatment and late post-treatment. However, there was a trend towards enrichment of IgA and increased persistent IgA clones, suggesting the survival of antigen experienced cells (**Supplementary Fig. 8b**).

### Transcriptomic changes in repopulated naïve B cells in rituximab responders

Following rituximab, substantial turnover in B cell populations occurred in responders. To investigate transcriptomic differences between depleted and repopulated B cells, we performed differential gene expression analysis between pretreatment and late post-treatment in responders. We focused specifically on naïve B cells as these were completely depleted at early post-treatment (**Supplementary Fig. 9**) and all patients had at least 10 cells at both timepoints. Other B cell subtypes had insufficient cell numbers and were therefore excluded.

A total of 171 genes (60 upregulated and 111 downregulated) showed significant changes (FDR < 0.05, **Fig. 5a, Supplementary Table 3)**. Many genes, including *SOCS1*, *SOCS3*, *PTGER4*, *CD83*, *PRDM1*, *AIF1*, were related to the regulation of immune processes and have previously been reported in SLE GWAS (GWAS Catalog^29^: trait ‘systemic lupus erythematosus’). Pathway analysis further highlighted NF-κB pathway and GPCR signaling (**Fig. 5b**).

**Figure 5.**
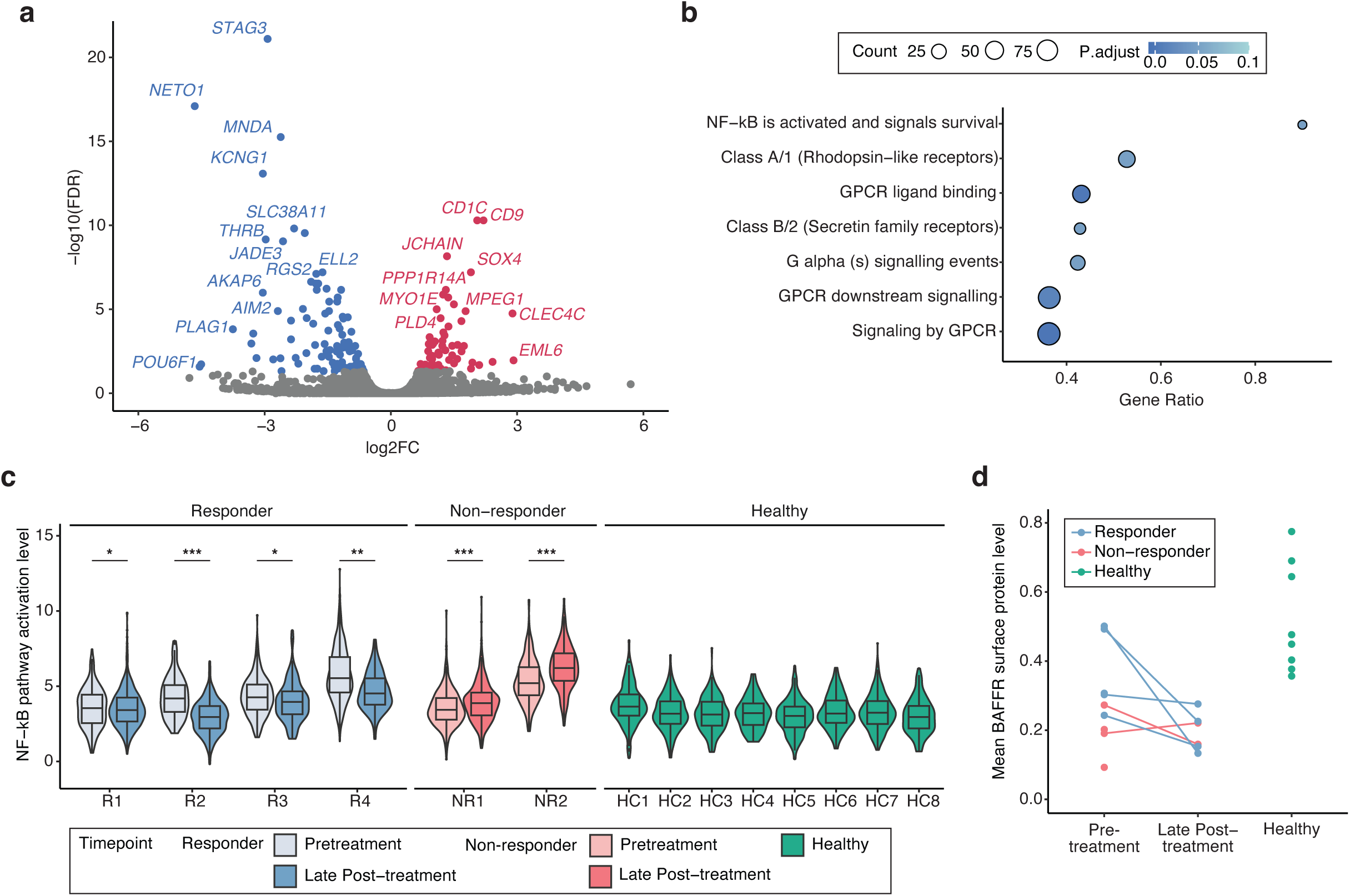
The transcriptomic profile of repopulated B cells in rituximab responders. **a)** Volcano plot showing DEGs (n = 171, FDR < 0.05) in naïve B cells from rituximab responders, comparing pretreatment to late post-treatment. Each point represents a gene, with significance and log_2_FC plotted on the y- and x-axes, respectively. **b)** Top Reactome pathways enriched among DEGs in responder naïve B cells between pretreatment and late post-treatment. Gene ratio indicates the proportion of core enrichment genes relative to the total number of genes within each pathway. **c)** NF-κB pathway activation scores in naïve B cells from individual patients at pretreatment and late post-treatment. Violin plots show the distribution of activation scores across cells, while box plots indicate the interquartile range (box), median (center line), and minimum/maximum values excluding outliers (whiskers). Differences in pathway activation between time points were assessed per patient using the Wilcoxon rank-sum test. Adjusted p-values: *L<L0.05; **L<L0.01; ***L<L0.001. **d)** Mean BAFF-R cell surface protein levels on naïve B cells from healthy controls and SLE patients at pretreatment and late post-treatment.

Given the central role of NF-κB signaling in immune regulation, we sought to quantify its activity at the single-cell level. We calculated the NF-κB pathway activation level of each individual cell (**Supplementary Methods**). In all responders, the level of NF-κB pathway activation was significantly lower in late post-treatment naïve B cells compared to pretreatment (Wilcoxon rank-sum test p-value < 0.05), resembling levels in healthy controls (**Fig. 5c)**. Non-responders had higher activation levels at late post-treatment. A similar pattern was also observed in memory B cell subtypes, including ABCs (**Supplementary Fig. 10a**).

B-cell activating factor receptor (BAFF-R), when in contact with BAFF, activates the NF-κB pathway^30^. Hence, BAFF-R cell surface protein levels on naïve B cells were investigated to assess whether changes in receptor expression could be linked to altered NF-κB pathway activity (**Fig. 5d, Supplementary Fig. 10b**). BAFF-R surface protein levels were significantly lower at late post-treatment compared to pretreatment in 3/4 responders, aligning with the change in NF-κB pathway activation.

### Gene expression changes interacting with rituximab response in non-B cells

Broader immune responses may differ between responders and non-responders following rituximab. To investigate this, we analyzed gene expression changes in non-B cells by comparing response-specific effects from pretreatment to early post-treatment (**Methods**). For instance, this approach identifies genes that increase in expression in responders while decreasing in non-responders.

A total of 99 genes (72 unique) in 9 immune cell subtypes showed differential expression changes based on rituximab response (FDR < 0.05, **Fig. 6a, Supplementary Table 4**). Among these, CD4 TCMs and DN T cells had the highest number of significant genes, with 29 and 15, respectively. From permutation analysis, this is greater than expected by chance (empirical p-value <0.05, **Supplementary Fig. 11**).

**Figure 6.**
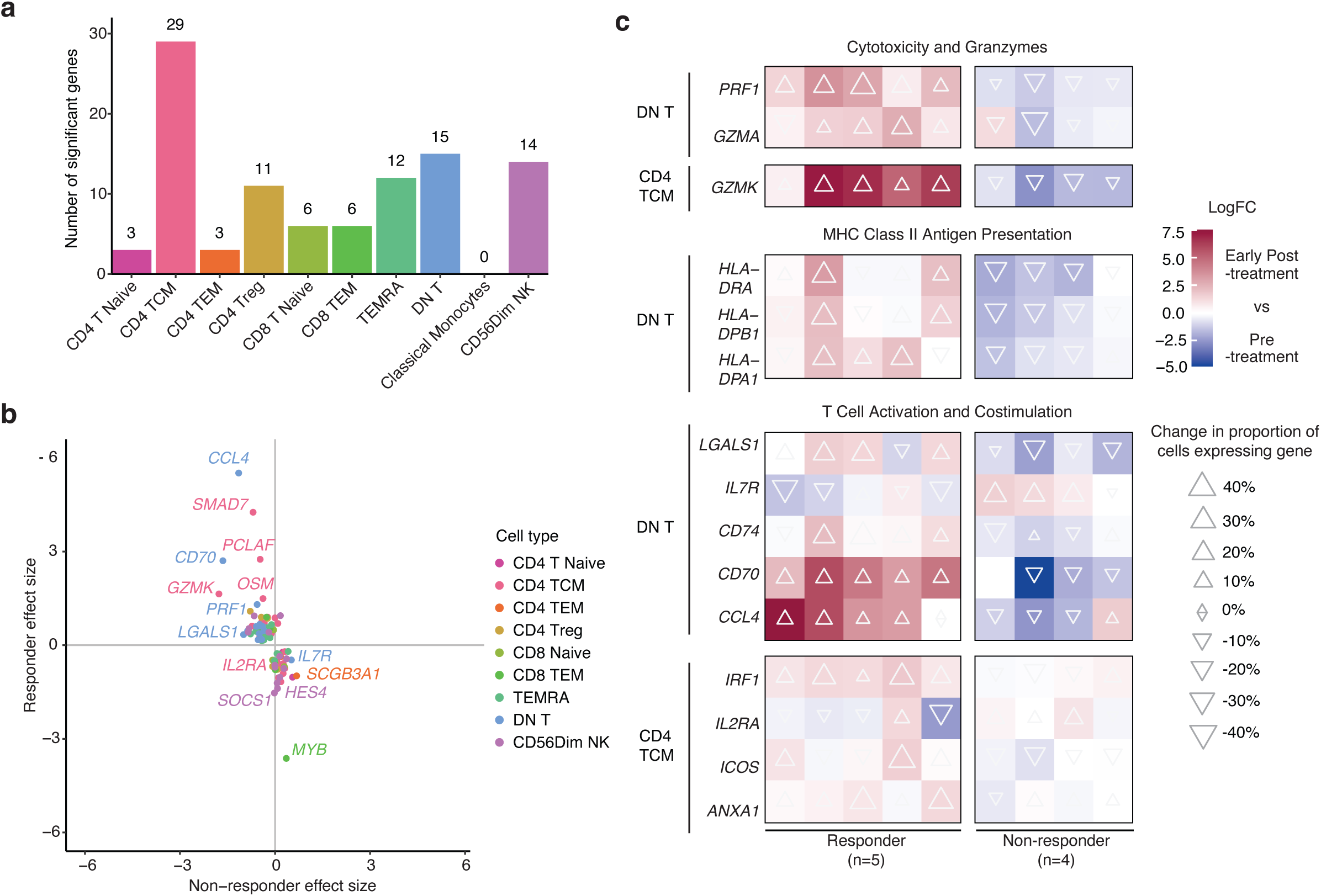
Gene expression changes interacting with rituximab response in non-B cells. **a)** The number of significant genes in non-B cell subtypes where rituximab response status alters gene expression changes between pretreatment and early post-treatment (FDR<0.05). Only cell types with at least 10 cells in all patients at both time points were analyzed. **b)** Effect sizes of significant genes where response status modifies gene expression changes between pretreatment and early post-treatment. The x-axis shows the effect size in non-responders, and the y-axis shows the effect size in responders. Each point represents a gene and is colored by the immune cell subtype in which it was significantly altered. **c)** Significant genes in DN T cells and CD4 TCM cells which function in ‘cytotoxicity and granzymes’, ‘MHC class II antigen presentation’ and ‘T cell activation and costimulation’. The fold changes in expression are shown as color, and changes in the proportion of cells expressing the gene are shown in triangle size and direction. Log_2_FC was calculated from TMM normalized expression levels. Expression values were not adjusted for covariates that were included in the model for testing differential gene expression (patient and cellular detection rate). Gene function annotation was informed by Gene Ontology.

Of the 99 genes, 63 showed a greater, positive effect size in responders, while non-responders showed smaller, negative changes (**Fig. 6b**). Many DEGs were involved in cell cytotoxicity (*PRF1, GZMA, GZMK*), MHC class II antigen presentation (*HLA-DRA, HLA-DPB1, HLA-DPA1*), and T cell activation (*LGALS1, IL7R, CD74, CD70, CCL4, IRF1, IL2RA, ICOS, ANXA1*). Most of these genes had increased expression and a higher proportion of expressing cells in responders (**Fig. 6c**). This suggested distinct transcriptomic changes in non-B cells depending on clinical response.

## Discussion

Through single-cell profiling of immune cells, our longitudinal cohort revealed changes in immune cell dynamics following rituximab treatment in SLE. Rituximab led to significant early depletion of naïve and memory B cells, followed by repopulation dominated by naïve and transitional B cells. As a result, rituximab also affected the BCR repertoire, with sequences at early post-treatment being more mutated, class-switched and clonally expanded, as previously reported^12,31^. This contrasts with changes in the repertoire observed after CD19 CAR-T cell therapy, where IgG and IgA expressing cells are almost completely depleted^11^. Persistent B cell clones could also be identified, and these were enriched for more mature memory and antibody secreting cells. This expands on previous findings in patients with SLE treated with rituximab^12^, and these clones share similar features to persistent clones identified in patients with myasthenia gravis receiving rituximab^32^. Overall, changes in the repertoire were not sustained over time, returning to baseline by late post-treatment.

We also observed differences in IGHJ usage and CDR3 length prior to treatment between SLE patients and healthy controls that were specific to naïve and transitional cells (**Fig. 4f, g**). Biased VDJ usage, including for IGHJ genes, has been previously reported in SLE^33^. Certain V genes, including IGHV4-34, have also been implicated in SLE pathology^34^, but we found no significant differences in use of these genes. Both longer and shorter CDR3 lengths have also been reported in SLE^12,35,36^. Long CDR3 regions are associated with autoreactivity and are normally selected against during development^37^. These conflicting findings may be due to the cell populations sampled, with shorter CDR3 being reported in total B cells or naïve B cells, as in this study.

A small number of genes exhibited differential expression in non-B cells post-rituximab. Certain interferon pathway genes decreased in Treg, naïve CD8 T, effector memory CD8 T, DN T and CD56Dim NK cells, but not globally. This contrasts with the broad decrease in interferon pathway genes in T cells and monocytes found in CAR-T cell mediated B cell depletion therapy^11^. This may be due to the difference in the target cells (CD20^+^ vs CD19^+^), as antibody secreting cells (CD20^-^CD19^+^ plasma cells and plasmablasts) that remain after rituximab may contribute to the interferon pathway activation in non-B cells. Also, CAR-T therapy may lead to greater B cell depletion in lymphoid organs affecting a broader range of immune cells.

Using single-cell data, we were able to explore specific memory B cell subtypes, including ABCs. ABCs are a unique population of memory B cells that potentially contribute to autoimmune diseases by developing through TLR7/9 stimulation and produce autoantibodies and inflammatory cytokines in autoimmune diseases^38,39^. In SLE, ABC frequency correlates with disease activity^38,40^, and their deletion in MRL.lpr lupus-prone mice suppresses disease^41^. A previous study demonstrated a significant reduction of ABCs (CD11c^+^CD21^-^) following rituximab^42^. In our study, ABCs were fully depleted in responders and remained lowly abundant for an extended period even when overall B cell proportion in PBMC returned to pretreatment levels. In non-responders, we saw incomplete depletion in two out of four patients. As a disease-relevant cell type, further studies will be needed to clarify how the dynamics of ABC depletion contribute to response following rituximab treatment in SLE patients.

In our study, patients who clinically responded to rituximab showed increased abundance of transitional B cells at late post-treatment, while the memory B cell compartment remained lowly abundant. This finding aligns with previous research characterizing B cell repopulation after rituximab, where patients achieving full depletion had delayed repopulation and a B cell compartment primarily consisting of transitional B cells at 55 weeks^43^. Transitional B cells are immature precursors of naïve B cells and may play a regulatory role through production of IL-10 and other cytokines^44^. Therefore, this alteration in the balance between transitional B cells and memory B cells may reflect the response to rituximab. However, transitional B cells producing high levels of proinflammatory cytokines, such as IL-6^45^, have been associated with disease activity in SLE and could potentially contribute to relapse.

In non-B cell populations, several genes showed response-dependent transcriptional changes following rituximab. Notably, genes associated with T cell activation and cytotoxicity were upregulated in responder CD4 CTMs and DN T cells. In SLE, DN T cells are known to be expanded and contribute to disease pathology by promoting autoantibody production, secreting cytokines such as IL-4, IL-17, and IFN-γ, and infiltrating target organs like the kidney^46,47^. Interestingly, DN T cells have also been reported to regulate B cell apoptosis and inhibit B cell proliferation and plasma cell differentiation^48^. Our analysis identified increased expression of *LGALS1* in responder DN T cells and decreased expression in non-responders following treatment. This gene has been shown to regulate B cell apoptosis and has been associated with rituximab resistance in non-Hodgkin lymphoma^49^. These findings suggest that DN T cells may contribute to B cell apoptosis during rituximab-induced depletion.

Limitations to our study include the sample size and sample collection timepoints. A larger cohort would enable more comprehensive comparisons between responders and non-responders at each timepoint, including cell type proportions and gene expression. Additional time points would provide further insight into specific response trajectories.

By conducting longitudinal single-cell sequencing, our study provides valuable insights into cellular changes from rituximab-induced B cell depletion and highlights the role of various immune cells in SLE pathogenesis.

## Acknowledgements

We thank James E. Peters for his valuable clinical insights and feedback, Racheal Bashford for her assistance with BCR/TCR library preparation optimization, and Niek de Klein for his input on data quality control. We also acknowledge the use of ChatGPT (OpenAI) for writing support (rephrasing, improving grammar and clarity) during the preparation of this manuscript. We thank the patients and healthy volunteers who participated in the study and the support of the Community Partners of the Immunology Theme of the National Institute for Health and Care Research (NIHR) Imperial Biomedical Research Centre. We acknowledge the support of the health professionals in the Imperial Lupus Centre, Imperial College Healthcare NHS Trust. This research was supported by the National Institute for Health and Care Research (NIHR) Imperial Biomedical Research Centre. The views expressed are those of the authors and not necessarily those of the NIHR or the Department of Health and Social Care.

## Author Contributions

M.C.P., M.B., and E.E.D. conceived and designed the study. N.B. and T.S.R. performed the experiments. L.O. and K.L.B. provided key resources. M.C.P. was responsible for clinical data curation. H.J., C.S., W.L., and N.B. performed the formal analysis. H.J. and C.S. wrote the original draft of the manuscript. M.C.P., M.B., and E.E.D. supervised the project. All authors reviewed the manuscript.

## Data availability

The RNA-sequencing, CITE-seq and TCR and BCR sequencing will be available on EGA after publication.

## References

1. Dörner, T., Giesecke, C. & Lipsky, P. E. Mechanisms of B cell autoimmunity in SLE. Arthritis Res. Ther. 13, 243 (2011).

2. Zavaleta-Monestel, E. et al. Advances in systemic lupus erythematosus treatment with monoclonal antibodies: A mini-review. Cureus 16, e64090 (2024).

3. Rydén-Aulin, M. et al. Off-label use of rituximab for systemic lupus erythematosus in Europe. Lupus Sci Med 3, e000163 (2016).

4. Merrill, J. T. et al. Efficacy and safety of rituximab in moderately-to-severely active systemic lupus erythematosus: the randomized, double-blind, phase II/III systemic lupus erythematosus evaluation of rituximab trial. Arthritis Rheum. 62, 222–233 (2010).

5. Rovin, B. H. et al. Efficacy and safety of rituximab in patients with active proliferative lupus nephritis: the Lupus Nephritis Assessment with Rituximab study. Arthritis Rheum. 64, 1215–1226 (2012).

6. Thatayatikom, A. & White, A. J. Rituximab: a promising therapy in systemic lupus erythematosus. Autoimmun. Rev. 5, 18–24 (2006).

7. Furie, R. A. et al. B-cell depletion with obinutuzumab for the treatment of proliferative lupus nephritis: a randomised, double-blind, placebo-controlled trial. Ann. Rheum. Dis. 81, 100–107 (2022).

8. Reddy, V., Martinez, L., Isenberg, D. A., Leandro, M. J. & Cambridge, G. Pragmatic Treatment of Patients With Systemic Lupus Erythematosus With Rituximab: Long-Term Effects on Serum Immunoglobulins. Arthritis Care Res. 69, 857–866 (2017).

9. Robinson, J. I. et al. Comprehensive genetic and functional analyses of Fc gamma receptors influence on response to rituximab therapy for autoimmunity. EBioMedicine 86, 104343 (2022).

10. Carter, L. M. et al. Gene expression and autoantibody analysis reveals distinct ancestry-specific profiles associated with response to rituximab in refractory systemic lupus erythematosus. Arthritis Rheumatol (2022) doi:10.1002/art.42404.

11. Wilhelm, A. et al. Selective CAR T cell–mediated B cell depletion suppresses IFN signature in SLE. JCI Insight 9, (6 2024).

12. Bashford-Rogers, R. J. M. et al. Analysis of the B cell receptor repertoire in six immune-mediated diseases. Nature 574, 122–126 (2019).

13. Vander Heiden, J. A., et al. pRESTO: a toolkit for processing high-throughput sequencing raw reads of lymphocyte receptor repertoires. Bioinformatics 30, 1930–1932 (2014).

14. Ye, J., Ma, N., Madden, T. L. & Ostell, J. M. IgBLAST: an immunoglobulin variable domain sequence analysis tool. Nucleic Acids Res. 41, W34–40 (2013).

15. Gayoso, A. et al. Joint probabilistic modeling of single-cell multi-omic data with totalVI. Nat. Methods 18, 272–282 (2021).

16. Traag, V. A., Waltman, L. & van Eck, N. J. From Louvain to Leiden: guaranteeing well-connected communities. Sci. Rep. 9, 5233 (2019).

17. Dann, E., Henderson, N. C., Teichmann, S. A., Morgan, M. D. & Marioni, J. C. Differential abundance testing on single-cell data using k-nearest neighbor graphs. Nat. Biotechnol. 40, 245–253 (2022).

18. Sethna, Z., Elhanati, Y., Callan, C. G., Walczak, A. M. & Mora, T. OLGA: fast computation of generation probabilities of B- and T-cell receptor amino acid sequences and motifs. Bioinformatics 35, 2974–2981 (2019).

19. G. James, R., J. Ellison, C. & P. Crutchfield, J. dit: a Python package for discrete information theory. J. Open Source Softw. 3, 738 (2018).

20. Gupta, N. T. et al. Change-O: a toolkit for analyzing large-scale B cell immunoglobulin repertoire sequencing data. Bioinformatics 31, 3356–3358 (2015).

21. Robinson, M. D., McCarthy, D. J. & Smyth, G. K. edgeR: a Bioconductor package for differential expression analysis of digital gene expression data. Bioinformatics 26, 139–140 (2010).

22. Soneson, C. & Robinson, M. D. Bias, robustness and scalability in single-cell differential expression analysis. Nat. Methods 15, 255–261 (2018).

23. Wolf, F. A., Angerer, P. & Theis, F. J. SCANPY: large-scale single-cell gene expression data analysis. Genome Biol. 19, 15 (2018).

24. Milacic, M. et al. The Reactome Pathway Knowledgebase 2024. Nucleic Acids Res. 52, D672–D678 (2024).

25. Schubert, M. et al. Perturbation-response genes reveal signalling footprints in cancer gene expression. Nat. Commun. 9, 20 (2018).

26. Badia-I-Mompel, P., et al. decoupleR: ensemble of computational methods to infer biological activities from omics data. Bioinform Adv 2, vbac016 (2022).

27. Sims, G. P. et al. Identification and characterization of circulating human transitional B cells. Blood 105, 4390–4398 (2005).

28. Rönnblom, L. & Leonard, D. Interferon pathway in SLE: one key to unlocking the mystery of the disease. Lupus Sci. Med. 6, e000270 (2019).

29. Sollis, E. et al. The NHGRI-EBI GWAS Catalog: knowledgebase and deposition resource. Nucleic Acids Res. 51, D977–D985 (2023).

30. Gardam, S. & Brink, R. Non-canonical NF-κB signalling initiated by BAFF influences B cell biology at multiple junctures. Front. Immunol. 4, 509 (2014).

31. Rouzière, A.-S., Kneitz, C., Palanichamy, A., Dörner, T. & Tony, H.-P. Regeneration of the immunoglobulin heavy-chain repertoire after transient B-cell depletion with an anti-CD20 antibody. Arthritis Res. Ther. 7, R714 (2005).

32. Jiang, R., et al. Single-cell repertoire tracing identifies rituximab-resistant B cells during myasthenia gravis relapses. JCI Insight 5, (2020).

33. Ota, M. et al. Multimodal repertoire analysis unveils B cell biology in immune-mediated diseases. Ann. Rheum. Dis. 82, 1455–1463 (2023).

34. Richardson, C. et al. Molecular basis of 9G4 B cell autoreactivity in human systemic lupus erythematosus. J. Immunol. 191, 4926–4939 (2013).

35. Liu, S. et al. Direct measurement of B-cell receptor repertoire’s composition and variation in systemic lupus erythematosus. Genes Immun. 18, 22–27 (2017).

36. Yurasov, S. et al. Defective B cell tolerance checkpoints in systemic lupus erythematosus. J. Exp. Med. 201, 703–711 (2005).

37. Meffre, E. et al. Immunoglobulin heavy chain expression shapes the B cell receptor repertoire in human B cell development. J. Clin. Invest. 108, 879 (2001).

38. Jenks, S. A. et al. Distinct Effector B Cells Induced by Unregulated Toll-like Receptor 7 Contribute to Pathogenic Responses in Systemic Lupus Erythematosus. Immunity 49, 725–739.e6 (2018).

39. Mouat, I. C., Goldberg, E. & Horwitz, M. S. Age-associated B cells in autoimmune diseases. Cell. Mol. Life Sci. 79, 402 (2022).

40. Jacobi, A. M. et al. Activated memory B cell subsets correlate with disease activity in systemic lupus erythematosus: delineation by expression of CD27, IgD, and CD95. Arthritis Rheum. 58, 1762–1773 (2008).

41. Nickerson, K. M. et al. Age-associated B cells are heterogeneous and dynamic drivers of autoimmunity in mice. J. Exp. Med. 220, (2023).

42. Faustini, F. et al. Rituximab in Systemic Lupus Erythematosus: Transient Effects on Autoimmunity Associated Lymphocyte Phenotypes and Implications for Immunogenicity. Front. Immunol. 13, 826152 (2022).

43. Sutter, J. A. et al. A longitudinal analysis of SLE patients treated with rituximab (anti-CD20): factors associated with B lymphocyte recovery. Clin. Immunol. 126, 282–290 (2008).

44. Nova-Lamperti, E. et al. IL-10-produced by human transitional B-cells down-regulates CD86 expression on B-cells leading to inhibition of CD4+T-cell responses. Sci. Rep. 6, 20044 (2016).

45. Liu, M. et al. Type I interferons promote the survival and proinflammatory properties of transitional B cells in systemic lupus erythematosus patients. Cell. Mol. Immunol. 16, 367–379 (2019).

46. Crispín, J. C. et al. Expanded double negative T cells in patients with systemic lupus erythematosus produce IL-17 and infiltrate the kidneys. J. Immunol. 181, 8761–8766 (2008).

47. Poddighe, D., Dossybayeva, K., Kozhakhmetov, S., Rozenson, R. & Assylbekova, M. Double-Negative T (DNT) Cells in Patients with Systemic Lupus Erythematosus. Biomedicines 12, (2024).

48. Hu, S.-H. et al. NKG2D Enhances Double-Negative T Cell Regulation of B Cells. Front. Immunol. 12, 650788 (2021).

49. Lykken, J. M. et al. Galectin-1 drives lymphoma CD20 immunotherapy resistance: validation of a preclinical system to identify resistance mechanisms. Blood 127, 1886–1895 (2016).

