## Supplementary methods for "Single-cell level characterization of B cell depletion and repopulation following rituximab in systemic lupus erythematosus"

Treatment Response

Response to rituximab was defined by a clinician at 12 months post-treatment. Patients were classified as responders if clinical SLEDAI (cSLEDAI: modified SLE Disease Activity Index 2000 which does not take serology into account^1^ was ≤ 2 and required ≤ 4mg prednisolone once daily at 12 months. When the sole manifestation was proteinuria, patients were classified as responders when the patient was non-nephrotic with >50% reduction in urine protein : creatinine ratio (uPCR).

Sample collection

A total of 40 samples were collected: 9 pretreatment samples and 23 post-treatment samples from SLE patients and 8 healthy control samples. All 40 samples were included in the sequencing and QC to maximize the number of samples available for cell type annotation. For patients with more than one sample collected within a timepoint, the later sample was excluded from analysis (n=2). Additionally, 6 post-treatment samples following further biologic therapy were also excluded from analysis, leaving 32 samples for analysis.

PBMC isolation and cryopreservation

PBMCs were isolated through density gradient centrifugation using Lymphoprep (STEMCELL Technologies, Canada). Approximately 20 ml of blood were diluted 1× with phosphate buffered saline (PBS) with addition of 2% fetal bovine serum (FBS) and layered on top of 15 ml of Lymphoprep solution. The samples were centrifuged at 800g for 20 minutes at room temperature without a break. PBMCs were collected from the interface and washed twice with PBS supplemented with 2% FBS. The cells were cryopreserved in 1 ml of freezing medium (FBS with 10% DMSO) and stored in liquid nitrogen.

Library preparation and sequencing

Cryopreserved peripheral blood mononuclear cells (PBMCs) were thawed, counted and resuspended for library preparation. Sample purification was done with a Stemcell EasySep™ Dead Cell Removal Kit (Catalog # 17899). For cellular indexing of transcriptomes and epitopes (CITE-seq), TotalSeq™-C Human Universal Cocktail V1.0 was used for antibody staining of 137 surface protein targets. Libraries were prepared according to the manufacturer's protocol.

10 pools were created with an equal number of cells from 4 samples each. Samples collected at different timepoints from the same patient were distributed across different pools to allow demultiplexing of individuals using genotypes captured from scRNA-seq data. Pools were then processed using the 10x Chromium platform for generation of gene expression (GEX), surface protein (CITE), TCR-enriched and BCR-enriched libraries. The Chromium Next GEM Single Cell 5’ Reagent Kit v2 (10x Genomics), Chromium Next GEM Chip K Single Cell Kit, Chromium Single Cell Human TCR Amplification Kit and Chromium Single Cell Human BCR Amplification Kit were used according to manufacturer’s instructions with 33,000 cells from each pool loaded onto 2 lanes of the chip.  Resulting libraries were pooled using a ratio of GEX:CITE:TCR:BCR=9:2:1:1 and were sequenced using NovaSeq 6000 (Illumina).

Bulk BCR repertoire sequencing

After scRNA-seq, the remaining PBMC samples were frozen in a lysis buffer. Reverse transcription was carried out using a pool of reverse primers specific to each immunoglobulin constant region and containing both a unique molecular identifier (UMI) and common tag region. Amplification by multiplex PCR was then performed using a set of primers targeting the FWR1 of the IGHV region and a reverse primer binding the common tag region^2^. Forward primers contained an internal barcode to allow for downstream sample demultiplexing. This was followed by library preparation and sequencing on Illumina MiSeq. The repertoire from one responder pretreatment sample could not be successfully amplified.

Single-cell data preprocessing and demultiplexing

Background droplets were removed, and sequencing reads were aligned to the GRCh38 human genome using Cell Ranger multi v7.0.0 (10x Genomics). The QC steps taken to remove low quality cells are outlined in **Supplementary Fig. 1**. First, cells with log transformed gene count or read count of lower than median - 3 x median absolute deviation were removed from each pool. Cells with mitochondrial read percentage, log-transformed CITE-seq feature counts or read counts greater than median + 3 x median absolute deviation were also removed from each pool, leaving 227,501 droplets after QC.

To assign cells to individuals by demultiplexing pools, we used genotypes identified from RNA reads in individual cells. cellSNP-lite(v.1.2.3)^3^ was first used to pile up expressed alleles, which was followed by Vireo(v.0.2.3)^4^ with default settings to allocate cells to respective individuals. 30,727 droplets classified as ‘doublets’ or ‘unassigned’ (due to low demultiplexing accuracy) were excluded from further analyses. With demultiplexed samples, we assessed genetic correlations using GenotypeMatcher^5^, and identified samples collected from the same individual.

Additionally, Souporcell (v.2.5)^6^ was used to evaluate the quality and accuracy of Vireo demultiplexing results. Although most lanes were demultiplexed uniformly between Vireo and Souporcell, there were two individuals in one of the sequencing lanes for pool 4 that exhibited different assignments, and a high number of cells were classified as doublets or unassigned cells. To ensure accurate demultiplexing, cells assigned to either individual from that lane were disregarded (5,821 droplets). However, the paired lane with identical samples loaded had high demultiplexing accuracy and was retained for subsequent analyses.

An additional 5,475 doublets were removed using VDJ information from Cell Ranger by identifying droplets with > 1 TRB chain, > 1 BCR heavy chain, or both TCR and BCR chains. Filtered BCR and TCR contigs from Cell Ranger were further processed using the Immcantation framework. V(D)J assignment was carried out using IgBLAST(v1.21.0)^7^ with AssignGenes.py and MakeDb.py from Change-O^8^. 13,918 and 67,062 BCR and TCR sequences from cells annotated as B cells and with a single paired heavy and light chain or annotated as T cells and with a single paired alpha and beta chain, were used in downstream repertoire analyses, respectively.

Single-cell data clustering and cluster level quality control

Gene expression values were log+1 normalized (Scanpy v1.10.1^9^: log1p) and the top 4,000 highly variable genes were identified using the Seurat v3 algorithm^10^. Multimodal integration and batch correction were performed with TotalVI (scvi-tools v1.1.2^11^) on default settings with all 137 proteins and highly variable genes. 20 low-dimensional latent representations inferred by totalVI were then used for neighborhood construction (k=15) and generation of uniform manifold approximation and projection (UMAP) for visualization. Leiden algorithm^12^ was used for unsupervised clustering with resolution=2 for annotation.

In addition to doublets identified by Vireo, we also used Scrublet (v0.2.3)^13^ and DoubletDetection(v3.0)^14^ to identify clusters of doublets consisting of cells of the same genotype. After confirming that unique cell type markers from different cell types were simultaneously expressed in these clusters, we removed 4,664 doublets. Clusters with distinctively low ribosomal read percentage were also removed (10,817 cells from cluster 16, 28, 30, 31, 32, and 39 in **Supplementary Fig. 2**). These cells had higher mitochondrial read percentages and expression of lncRNAs acting as transcriptional regulators in the nucleus, suggesting ruptured cells. After removing these clusters, normalization, integration, and clustering was redone on the dataset to obtain the final UMAP (**Supplementary Fig. 3**).

Cell type annotation

We first used Azimuth^15^ (v0.5.0: Human PBMC reference) and Celltypist^16^ (v1.6.3: Healthy_COVID19_PBMC reference model) to conduct automated PBMC cell type annotation by label transfer. Clusters annotated as platelets and erythroid cells (484 cells) from both programs were removed as contaminants, likely resulting from incomplete centrifugation during PBMC extraction. Using the two automated annotations as guidelines, annotations were then further refined using denoised surface protein values and normalized gene expression values. The dataset was first split into T cells, B cells, and other immune cells and these subsets were re-clustered with the Leiden algorithm with resolution=1 to capture minor cell subtypes.

Preprocessing of bulk BCR repertoire data

Raw sequence data were initially processed using Presto^17^ from the Immcantation analysis suite. First, paired end reads were assembled and barcodes introduced by the PCR forward primer were used to demultiplex samples. Assembled reads with a mean Phred score under 20 were removed. UMIs introduced during reverse transcription were identified and reads with identical UMIs were collapsed into a single consensus sequence. VDJ assignment was then performed using IgBLAST v1.21.0 via the Change-O wrapper. IMGT reference sequences^18^ were downloaded 09/10/2023. Sequences with UMIs of unexpected length (other than 14 nucleotides) and containing internal N nucleotides were removed. An issue with amplification of IGHV4 was detected in samples processed using the same primer pool. To enable comparison between samples despite this technical issue, sequences with an IGHV4 gene assignment were dropped from all samples. In total, 1,028,618 unique sequences were retained.

Differential abundance analysis

We identified differentially abundant cell populations between timepoints (pretreatment vs. early post-treatment, early post-treatment vs. late post-treatment, and pretreatment vs. late post-treatment) using MiloR (v.2.0)^19^. Using the 20 batch-corrected latent dimensions from TotalVI, we constructed cell neighborhoods from the k-nearest neighbours graph (k=60). A mixed effect model adjusting for individual patient effects as a random intercept was used to test differential abundance within each neighborhood. Given the depletion of B cells at the early post-treatment timepoint, a pseudocount of 0.1 was added to each sample in each neighborhood to avoid inflation of fold change when cells were completely absent. neighborhoods were annotated as the cell type of most cells within the neighborhood. For B cells, differential abundance was tested within each response group. Spatial FDR was used to determine significance, since it weights each P-value by the reciprocal of the kth nearest neighbour distance, effectively accounting for the overlap between neighborhoods.

BCR and TCR repertoire analysis

When calculating repertoire metrics in the single cell BCR and TCR data, sequences from each unique cell were counted once. For the bulk BCR data, to account for varying numbers of antibody secreting cells, which have extremely high levels of BCR RNA^20^, each unique nucleotide sequence (as deduplicated using the CollapseSeq command from Presto) was counted once and analyses were not weighted by UMI counts. Shannon entropy was used for diversity calculations. This is a measure of uncertainty that reaches a maximum when a repertoire is composed of clonotypes of equal frequency. Bulk BCR samples were first size-matched by down-sampling to the same number of UMIs as present in the smallest sample (1660 UMIs). Shannon Entropy was then calculated on unique nucleotide junction regions (CDR3 plus the conserved C - 104 and W/F - 118 codons) using the *dit* Python package^21^. This process was repeated 1000 times and a mean calculated across these iterations. For the single cell TCR analysis, the same process was repeated but Shannon Entropy was calculated on unique clones (clones were defined as sequences sharing the same combination of alpha and beta amino acid junction region and VJ genes) and samples were down-sampled to 451 cells. Mutation frequency was defined as the number of bases differing from the germline sequence divided by the length of the sequence, with the D gene masked. Clonal grouping with Change-O was carried out with a normalized Hamming distance threshold of  0.1, established using the distToNearest function in SHazaM^22^*.* Statistical analyses were performed using SciPy^23^ , Pingouin^24^ or scikit-posthocs^25^ packages.

Differential gene expression analysis

To get pseudobulk gene expression values, we used Seurat2PB (Seurat v.5.0.3^26^) by sample and cell type. Batch correction was conducted using ComBat-seq (sva v3.50.0^27^) using samples from all individuals and timepoints. The timepoints of sample collection were used as a biological covariate to preserve their signals in the adjusted data. To reduce false positives, only genes with an estimated counts per million (CPM) above 1 in more than 25% of cells across all samples in comparison were retained. Genes on the Y chromosome, TCR/BCR genes, and haemoglobin genes (HBA1, HBA2, HBB) were also excluded from analysis. TMM (trimmed mean of M values) were used to calculate normalization factors with cell-type specific library sizes using edgeR(v.3.42.0)^28^ .

To understand the effect of potential covariates on overall gene expression, we applied VariancePartition(v.1.24.1)^29^ to SLE pretreatment samples and healthy control samples to calculate the variation explained by cell type, disease status, individual, cellular detection rate (the fraction of detected genes per cell to adjust for sequencing depth as described in Soneson et al^30^), age, and self-reported ancestry in pseudobulked gene expression. Age and self-reported ancestry contributed minimally to the variation found in the expression profile and were therefore excluded as covariates in the model. We applied the quasi-likelihood approach from edgeR^28^ to identify differentially expressed genes accounting for patient (to adjust for variance between patients) and cellular detection rate as covariates.

We conducted three comparisons to identify differentially expressed genes. To determine if there are differences between pretreatment cells and repopulated cells after treatment, we compared naive B cells from responders at pretreatment and late post-treatment for 10,491 highly expressed genes. In non-B cells, we first compared pretreatment to early post-treatment in all patients, regardless of response. Next, we identified genes in which response status alters the expression change between pretreatment and early post-treatment. We used the same pseudobulk approach described above, but we implemented the approach detailed in the edgeR User’s Guide (Section 3.5, version as of 29 April 2024) to identify genes where changes in expression between timepoints differed depending on treatment response. We first initiated the design matrix with patient effects and cellular detection rate, then defined response-specific time point effects and appended them to the design matrix. We then fit a linear model (glmQLFit) and contrasted the response-specific time point effects for early post-treatment to pretreatment. This allowed us to account for relatedness between the samples taken by the same patient in the comparison between timepoints.

To confirm our results were robust, a permutation analysis was conducted. We shuffled the response label for each patient to every permutation possible (126 cases) and calculated the number of significant genes observed. Significance of the differential gene expression analysis was defined using an empirical p-value less than 0.05. Informed by Gene Ontology (GO)^31^ gene lists, we grouped differentially expressed genes identified in CD4 TCM cells and DN T cells to ‘Cytotoxicity and Granzymes’(GO:0140507 and manual curation), ‘MHC Class II Antigen Presentation’ (GO:0002504), and ‘T Cell Activation and Costimulation’ (GO0042110, GO0031295).

NF-kB pathway activation scoring

Pathway activation scores were calculated for each cell using decoupleR(v.2.8.0)^32^ by fitting a univariate linear model between single-cell gene expression values and pathway-gene interaction weights for the top 500 responsive genes ranked by p-value in the NF-kB pathway in the PROGENy database. The obtained t-values of the slopes were used as activation scores. The NF-kB pathway activation scores and BAFFR surface protein levels were compared between pretreatment cells and late post-treatment cells within each patient using the Wilcoxon rank-sum test.

All preprocessing and analyses were conducted in R v4.1.3 or Python v3.11.

**References**

1.	Uribe, A. G. et al. The Systemic Lupus Activity Measure-revised, the Mexican Systemic Lupus Erythematosus Disease Activity Index (SLEDAI), and a modified SLEDAI-2K are adequate instruments to measure disease activity in systemic lupus erythematosus. J. Rheumatol. 31, 1934–1940 (2004).

2.	Bashford-Rogers, R. J. M. et al. Analysis of the B cell receptor repertoire in six immune-mediated diseases. Nature 574, 122–126 (2019).

3.	Huang, X. & Huang, Y. Cellsnp-lite: an efficient tool for genotyping single cells. Bioinformatics (2021) doi:10.1093/bioinformatics/btab358.

4.	Huang, Y., McCarthy, D. J. & Stegle, O. Vireo: Bayesian demultiplexing of pooled single-cell RNA-seq data without genotype reference. Genome Biol. 20, 273 (2019).

5.	de Klein, N. genotypeMatcher: Matching Vireo Genotypes between Pools. (Github).

6.	Heaton, H. et al. Souporcell: robust clustering of single-cell RNA-seq data by genotype without reference genotypes. Nat. Methods 17, 615–620 (2020).

7.	Ye, J., Ma, N., Madden, T. L. & Ostell, J. M. IgBLAST: an immunoglobulin variable domain sequence analysis tool. Nucleic Acids Res. 41, W34–40 (2013).

8.	Gupta, N. T. et al. Change-O: a toolkit for analyzing large-scale B cell immunoglobulin repertoire sequencing data. Bioinformatics 31, 3356–3358 (2015).

9.	Wolf, F. A., Angerer, P. & Theis, F. J. SCANPY: large-scale single-cell gene expression data analysis. Genome Biol. 19, 15 (2018).

10.	Stuart, T. et al. Comprehensive Integration of Single-Cell Data. Cell 177, 1888–1902.e21 (2019).

11.	Gayoso, A. et al. Joint probabilistic modeling of single-cell multi-omic data with totalVI. Nat. Methods 18, 272–282 (2021).

12.	Traag, V. A., Waltman, L. & van Eck, N. J. From Louvain to Leiden: guaranteeing well-connected communities. Sci. Rep. 9, 5233 (2019).

13.	Wolock, S. L., Lopez, R. & Klein, A. M. Scrublet: Computational Identification of Cell Doublets in Single-Cell Transcriptomic Data. Cell Syst 8, 281–291.e9 (2019).

14.	Shor, C. A. G. JonathanShor/DoubletDetection: Doubletdetection v4.2. doi:10.5281/zenodo.6349517.

15.	Hao, Y. et al. Integrated analysis of multimodal single-cell data. Cell 184, 3573–3587.e29 (2021).

16.	Domínguez Conde, C. et al. Cross-tissue immune cell analysis reveals tissue-specific features in humans. Science 376, eabl5197 (2022).

17.	Vander Heiden, J. A. et al. pRESTO: a toolkit for processing high-throughput sequencing raw reads of lymphocyte receptor repertoires. Bioinformatics 30, 1930–1932 (2014).

18.	Lefranc, M.-P. & Lefranc, G. The Immunoglobulin FactsBook. (Academic Press, San Diego, CA, 2001).

19.	Dann, E., Henderson, N. C., Teichmann, S. A., Morgan, M. D. & Marioni, J. C. Differential abundance testing on single-cell data using k-nearest neighbor graphs. Nat. Biotechnol. 40, 245–253 (2022).

20.	Turchaninova, M. A. et al. High-quality full-length immunoglobulin profiling with unique molecular barcoding. Nat. Protoc. 11, 1599–1616 (2016).

21.	G. James, R., J. Ellison, C. & P. Crutchfield, J. dit: a Python package for discrete information theory. J. Open Source Softw. 3, 738 (2018).

22.	Yaari, G. et al. Models of somatic hypermutation targeting and substitution based on synonymous mutations from high-throughput immunoglobulin sequencing data. Front. Immunol. 4, 358 (2013).

23.	Virtanen, P. et al. SciPy 1.0: fundamental algorithms for scientific computing in Python. Nat. Methods 17, 261–272 (2020).

24.	Vallat, R. Pingouin: statistics in Python. J. Open Source Softw. 3, 1026 (2018).

25.	Terpilowski, M. scikit-posthocs: Pairwise multiple comparison tests in Python. J. Open Source Softw. 4, 1169 (2019).

26.	Hao, Y. et al. Dictionary learning for integrative, multimodal and scalable single-cell analysis. Nat. Biotechnol. 42, 293–304 (2024).

27.	Zhang, Y., Parmigiani, G. & Johnson, W. E. ComBat-seq: batch effect adjustment for RNA-seq count data. NAR Genom Bioinform 2, lqaa078 (2020).

28.	Robinson, M. D., McCarthy, D. J. & Smyth, G. K. edgeR: a Bioconductor package for differential expression analysis of digital gene expression data. Bioinformatics 26, 139–140 (2010).

29.	Hoffman, G. E. & Schadt, E. E. variancePartition: interpreting drivers of variation in complex gene expression studies. BMC Bioinformatics 17, 483 (2016).

30.	Soneson, C. & Robinson, M. D. Bias, robustness and scalability in single-cell differential expression analysis. Nat. Methods 15, 255–261 (2018).

31.	Gene Ontology Consortium et al. The Gene Ontology knowledgebase in 2023. Genetics 224, (2023).

32.	Badia-I-Mompel, P. et al. decoupleR: ensemble of computational methods to infer biological activities from omics data. Bioinform Adv 2, vbac016 (2022).
