## Supplementary figures and tables for "Single-cell level characterization of B cell depletion and repopulation following rituximab in systemic lupus erythematosus"

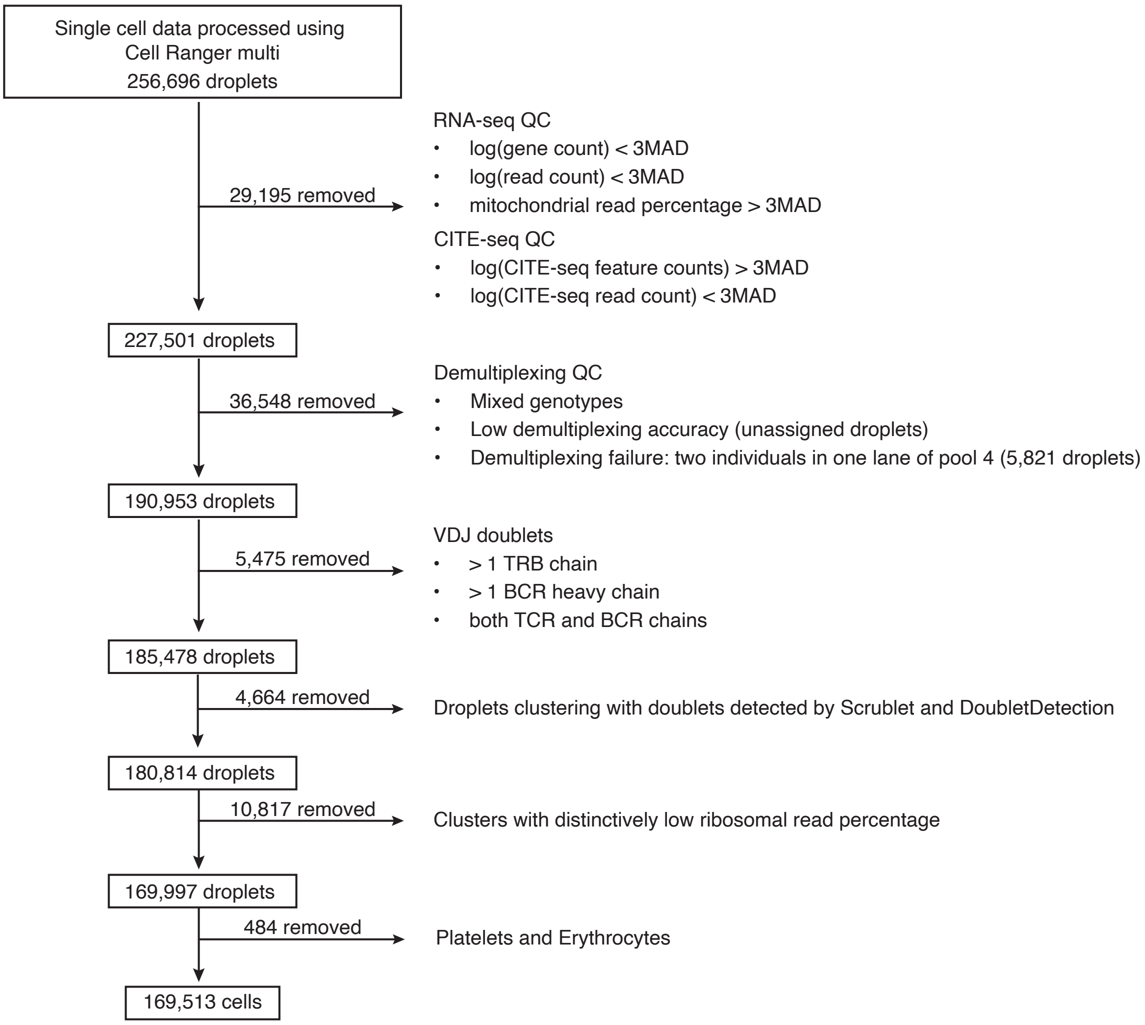


**Supplementary Figure 1.** Flowchart of the single-cell data quality control (QC) process, showing the number of droplets removed at each filtering step due to low quality. A total of 169,513 cells passed QC and were included in downstream analysis.

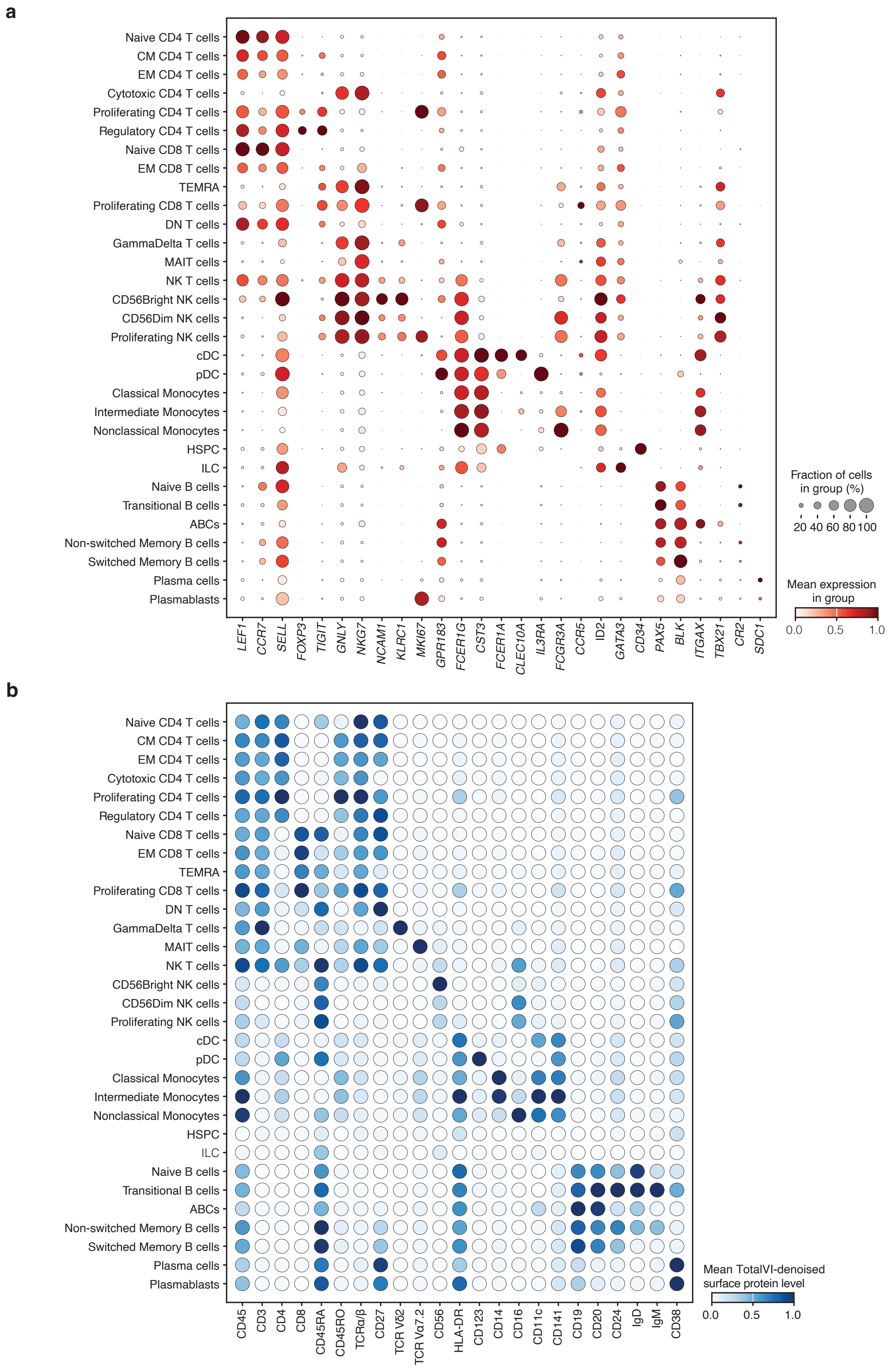


**Supplementary Figure 2.** Cell type markers used for immune cell annotation. **a)** Marker genes from scRNA-seq data used to annotate immune cell subtypes. Gene expression levels were normalized and log-transformed using Scanpy's default settings. **b)** Surface protein markers from CITE-seq data

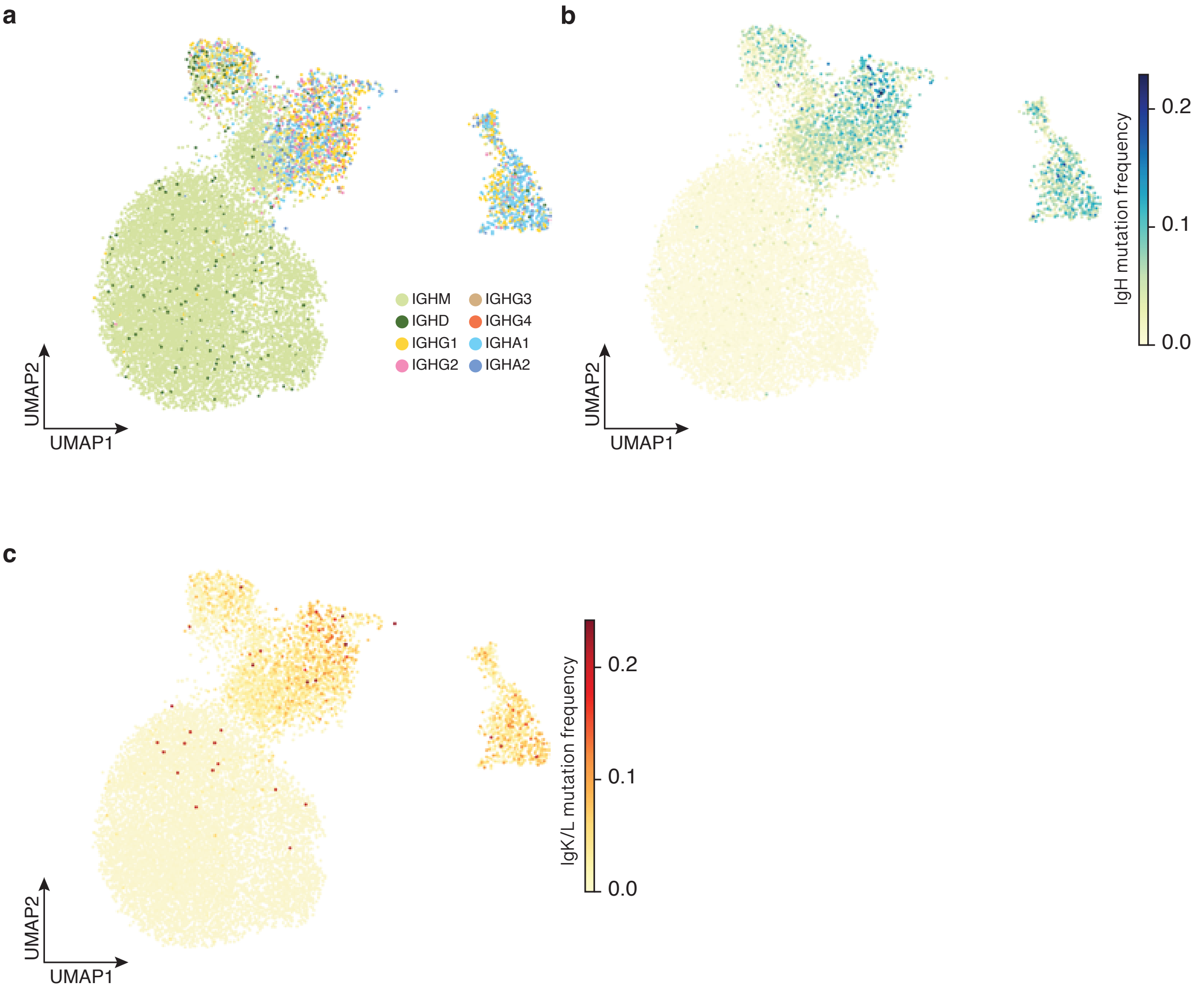


**Supplementary Figure 3. a)** Constant region gene usage from BCR data **b)** BCR heavy chain mutation frequency **c)** BCR light chain mutation frequency

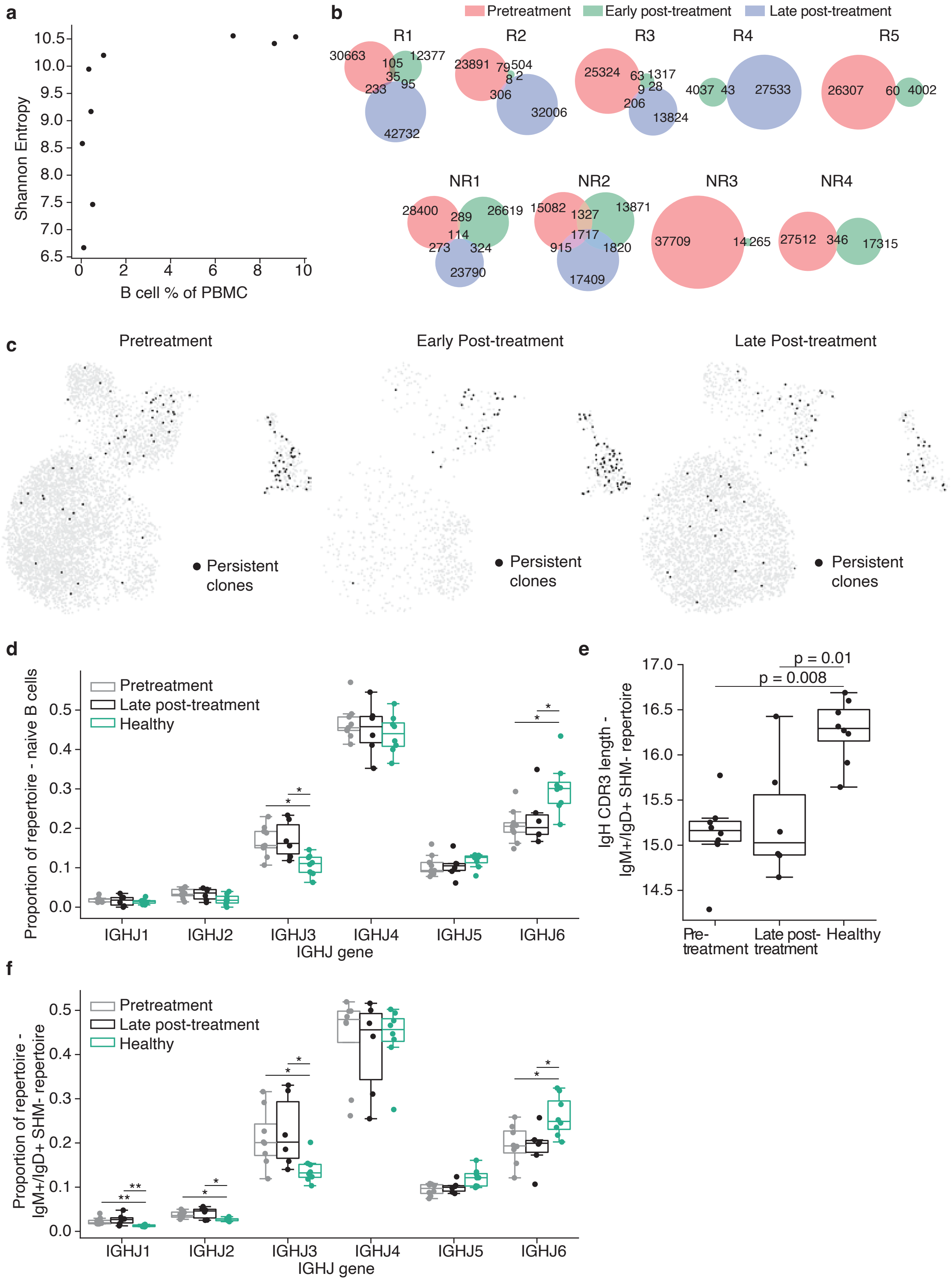


**Supplementary Figure 4. a)** Shannon Entropy of bulk repertoire against B cell % of PBMC from single cell data, both at early post-treatment. Higher values of Shannon Entropy indicate greater diversity. **b)** Numbers of unique and overlapping clones identified within each SLE patient across timepoints. **c)** B cell UMAP as shown in Fig. 2e highlighting cells with sequences belonging to persistent clones, split across timepoints. **d)** IGHJ gene usage in naïve B cells at pretreatment, late post-treatment and in healthy controls. Dunn's test was used for pairwise comparisons in genes with an FDR-adjusted p-value <0.05 from Kruskal-Wallis test. FDR-adjusted p-value *< 0.05. **e)** Mean IgH CDR3 length in IgM+/IgD+ SHM- sequences from bulk repertoire data. FDR adjusted p-value, Kruskal-Wallis followed by Dunn's test. **f)** IGHJ gene usage in IgM+/IgD+ SHM- sequences from bulk repertoire data at pretreatment, late post-treatment and in healthy controls. FDR-adjusted p-value *< 0.05 **< 0.01.

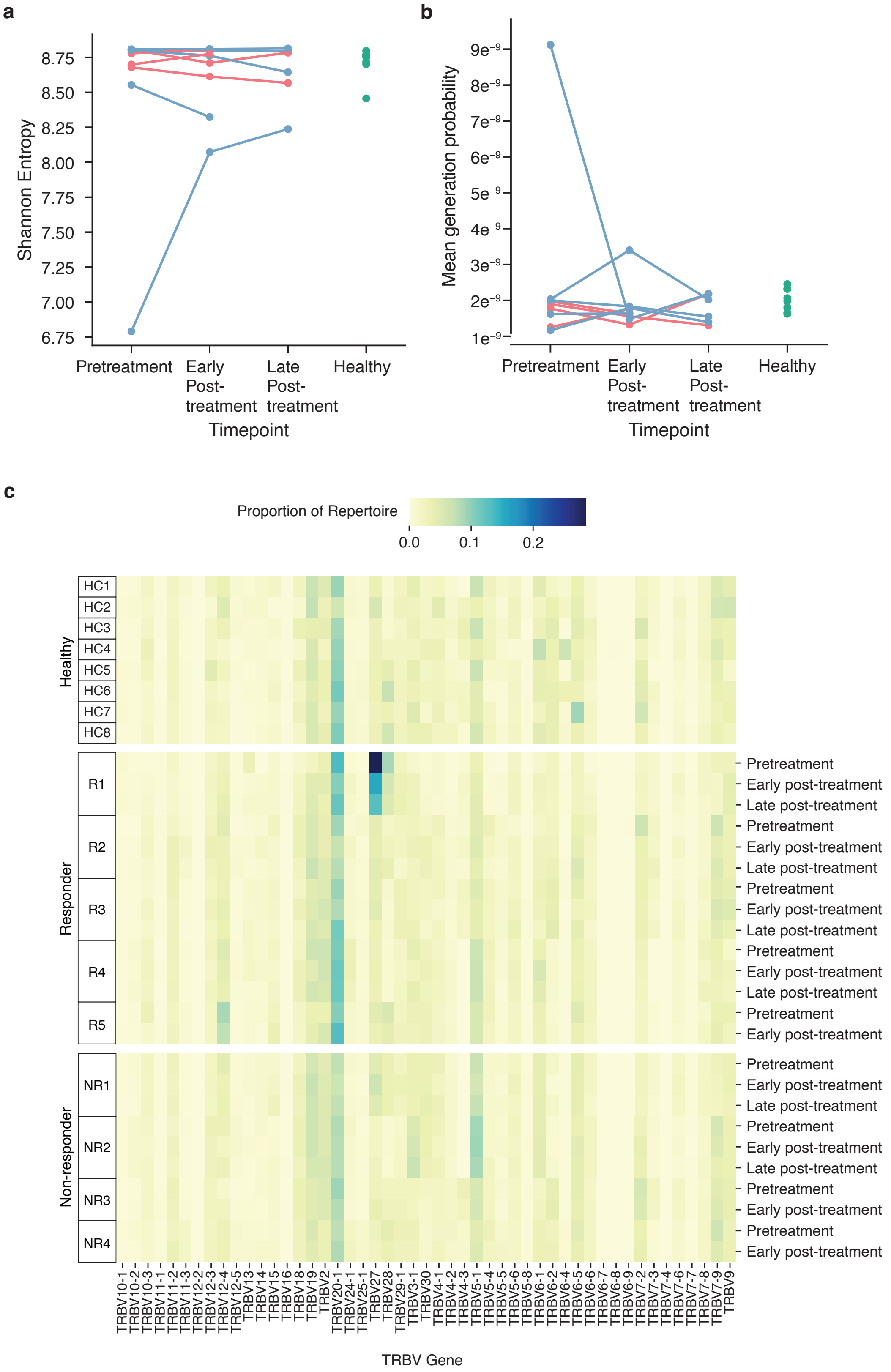


**Supplementary Figure 5. a)** Shannon Entropy of single cell TCR repertoire across timepoints. All samples were subset to 451 cells across 1000 iterations and a mean was calculated. **b)** Mean generation probability across timepoints, calculated by OLGA on nucleotide CDR3 sequence. Higher generation probability indicates a sequence that is more likely to occur by chance. **c)** Heatmap of TRBV gene usage across individuals and timepoints.

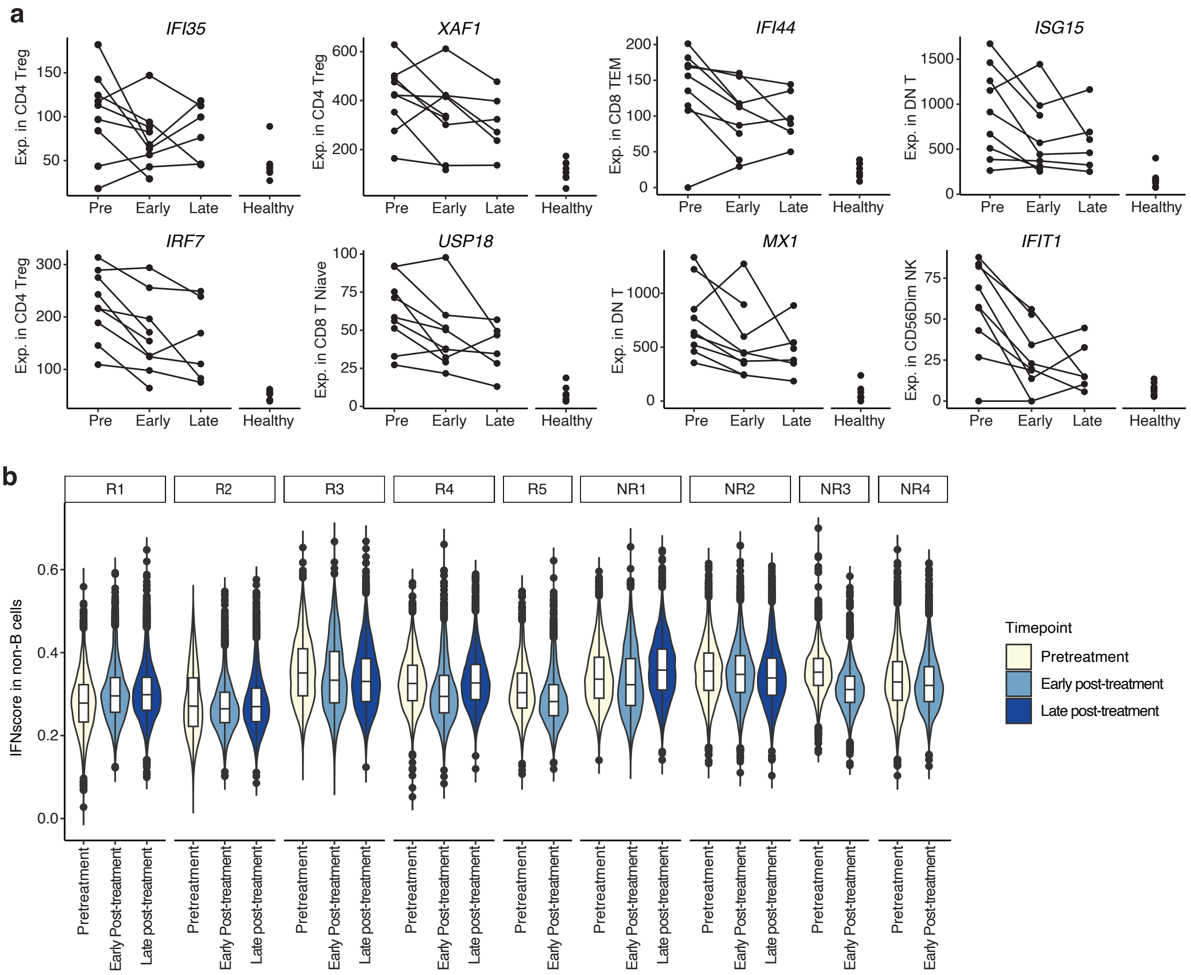


**Supplementary Figure 6. a)** Gene expression levels of interferon pathway genes that are significantly differentially expressed between pretreatment and early post-treatment in SLE patients. Gene expression levels were TMM normalised. **b)** Violin plot of interferon activity score in non-B cells from each patient and timepoint. Interferon score was calculated using genes from the Reactome gene set 'Interferon signaling (R-HSA-913531)'.

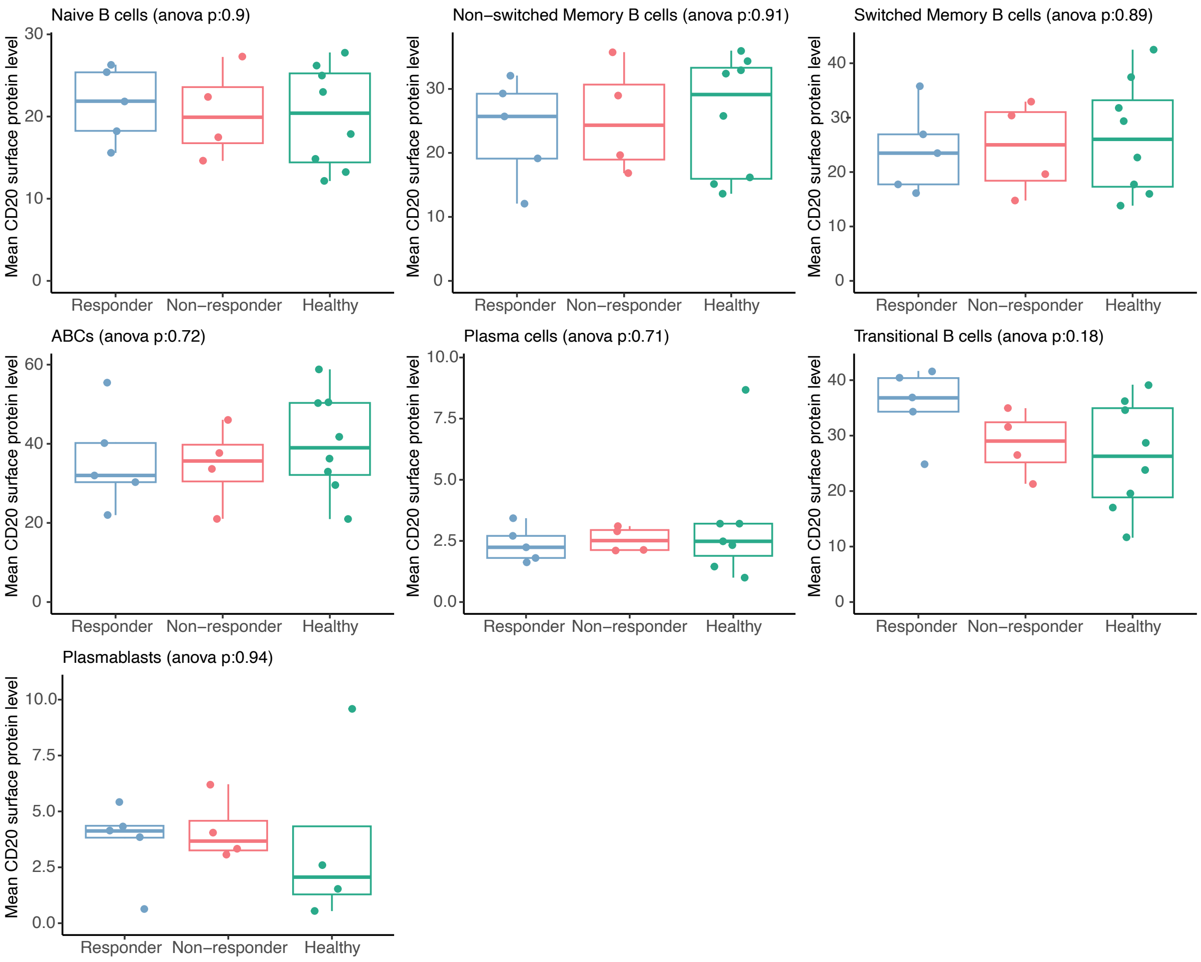


**Supplementary Figure 7.** CD20 surface protein levels of each B cell subtype at pretreatment in responders, non-responders and healthy controls. No significant differences were observed between groups with ANOVA (p-value > 0.05)

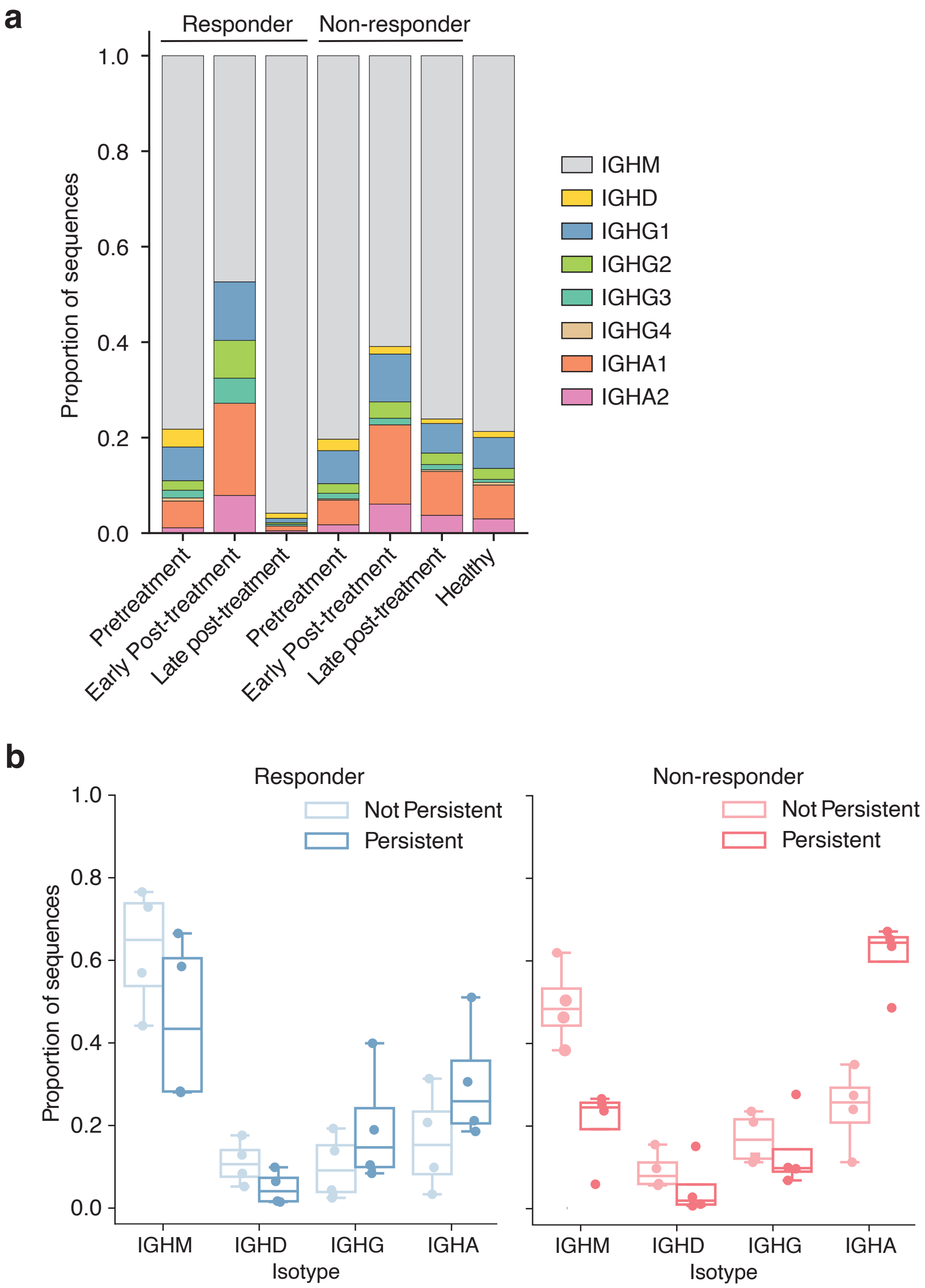


**Supplementary Figure 8. a)** Isotype usage in total B cells from single cell data, split by timepoint and rituximab response. **b)** Proportion of repertoire occupied by each isotype split between sequences forming persistent or non-persistent clones in responders and non-responders.

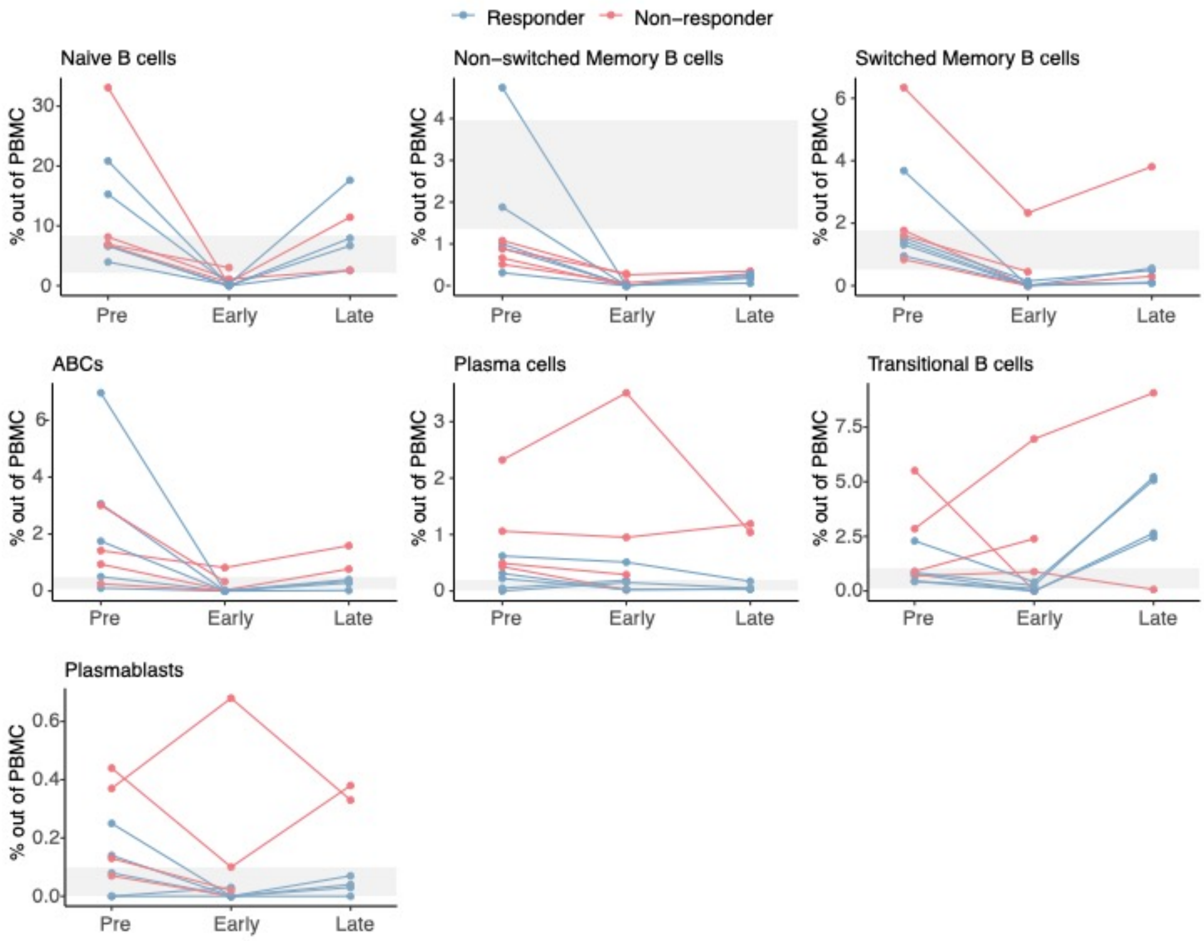


**Supplementary Figure 9.** The proportion of each B cell subtype out of all PBMC across timepoints by patient. scRNA-seq data was used to calculate cell type proportions. The shaded region shows the range of healthy controls. Pre = pretreatment; Early = early post-treatment; Late = late post-treatment

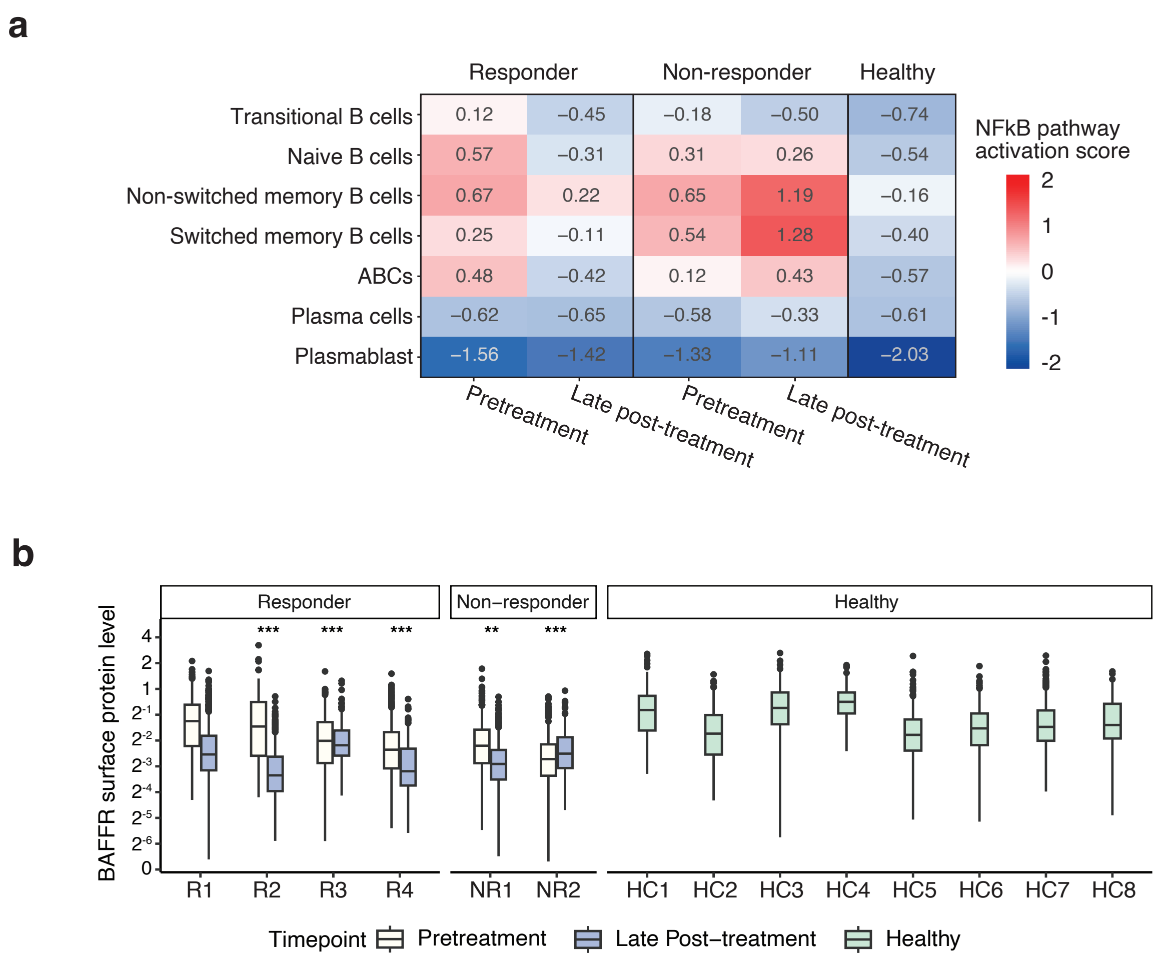


**Supplementary Figure 10. a)** Heatmap of mean NF-kB pathway activation score of B cell subtypes at pretreatment and late post-treatment. Score was calculated using decoupleR with PROGENy pathway gene weights and were scaled around zero. **b)** BAFFR cell surface protein levels on naive B cells. Two-sample Wilcoxon tests done between pretreatment and late post-treatment cells within each patient. *p-value < 0.05 **p-value < 0.01 ***p-value < 0.001

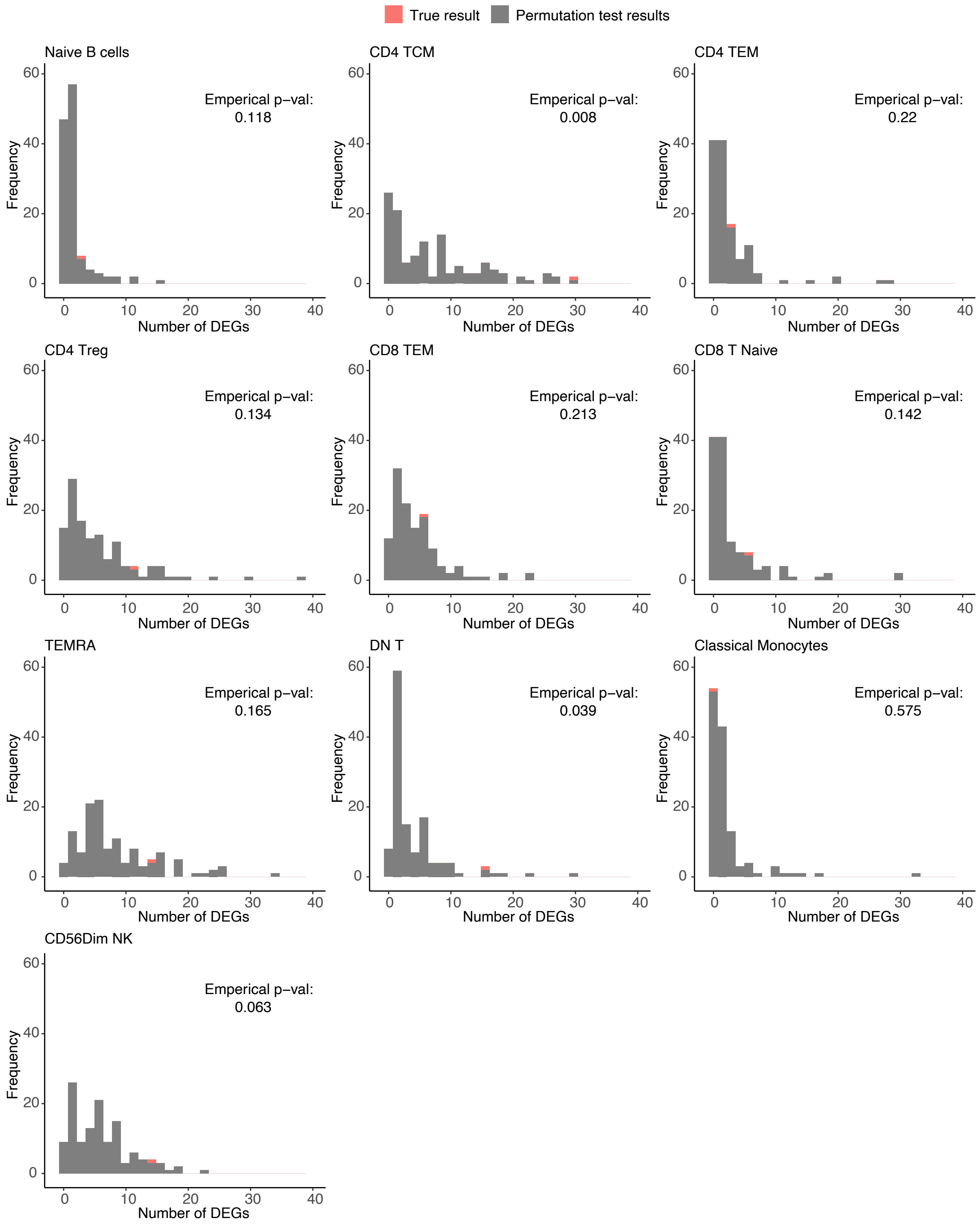


**Supplementary Figure 11.** Distribution of the number of significant genes from permutation analyses in non-B cells (FDR < 0.05). Genes in which response status alters the expression change between pretreatment and early post-treatment were tested while the response label for each patient was shuffled to all possible response permutations. The true result was considered significant if it had an empirical p-value < 0.05.

**Supplementary Table 1.** Participant demographics

| ID | Disease status | Age | Sex | Self-reported ancestry | Rituximab cycle | Lupus nephritis | dsDNA | ANA |
|---|---|---|---|---|---|---|---|---|
| R1 | SLE | 50-59 | F | Black Caribbean | 1 | Y | Negative | Positive |
| R2 | SLE | 20-29 | F | White Other | 1 | Y | Positive | Positive |
| R3 | SLE | 30-39 | F | White British | 1 | N | Positive | Positive |
| R4 | SLE | 30-39 | F | Black Caribbean | 1 | Y | Positive | Positive |
| R5 | SLE | 30-39 | F | White Other | 1 | Y | Positive | Positive |
| NR1 | SLE | 20-29 | F | Black Caribbean | 3 | N | Positive | Positive |
| NR2 | SLE | 30-39 | F | Black African | 1 | Y | Negative | Positive |
| NR3 | SLE | 50-59 | F | White Other | 1 | N | Positive | Positive |
| NR4 | SLE | 30-39 | M | Black Caribbean | 1 | Y | Positive | Positive |
| HC1 | Healthy | 30-39 | F | White British | | | | |
| HC2 | Healthy | 30-39 | F | White British | | | | |
| HC3 | Healthy | 30-39 | F | White Other | | | | |
| HC4 | Healthy | 30-39 | F | White British | | | | |
| HC5 | Healthy | 60-69 | F | White Other | | | | |
| HC6 | Healthy | 30-39 | F | Black African | | | | |
| HC7 | Healthy | 50-59 | F | White British | | | | |
| HC8 | Healthy | 50-59 | F | White Other | | | | |

**Supplementary Table 2.** Summary of clinical information at each sample collection timepoint for SLE patients

| ID | Rituximab Response[1] | Sample Time Point | Days from Rituximab | cSLEDAI[2] 2K | C3 | C4 | uPCR | eGFR | Lymphocyte Count | Neutrophil Count | MMF | Pred | HCQ | AZA | MTX |
|---|---|---|---|---|---|---|---|---|---|---|---|---|---|---|---|
| R1 | Responder | Pretreatment | 0 | 4 | 1.03 | 0.24 | 224 | 90 | 1.1 | 1.9 | Y | N | Y | N | N |
| R1 | Responder | Early post[2] | 70 | 4 | 1.39 | 0.33 | 539 | >90 | 0.8 | 2 | Y | N | Y | N | N |
| R1 | Responder | Late post[3] | 378 | 4 | 1.23 | 0.25 | 89 | 83 | 1.6 | 2.1 | Y | N | Y | N | N |
| R2 | Responder | Pretreatment | -30 | 4 | 0.68 | 0.14 | 174 | >90 | 1.2 | 2.7 | N | Y | Y | N | N |
| R2 | Responder | Early post | 81 | 0 | 0.83 | 0.18 | 53 | >90 | 1.5 | 3.9 | Y | N | Y | N | N |
| R2 | Responder | Late post | 431 | 0 | 0.85 | 0.18 | 18 | >90 | 1.3 | 3.2 | Y | N | Y | N | N |
| R3 | Responder | Pretreatment | 0 | 5 | 0.58 | 0.07 | 0 | >90 | 0.6 | 3.5 | N | Y | Y | N | N |
| R3 | Responder | Early post | 81 | 4 | 0.73 | 0.09 | 0 | >90 | 0.3 | 5.7 | N | Y | Y | N | N |
| R3 | Responder | Late post | 285 | 0 | 0.87 | 0.15 | 0 | >90 | 0.5 | 2.1 | N | Y | Y | N | N |
| R4 | Responder | Pretreatment | 0 | 4 | 0.19 | 0.03 | 151 | >90 | 0.3 | 2.7 | Y | Y | Y | N | N |
| R4 | Responder | Early post | 129 | 0 | 1.12 | 0.25 | 40 | >90 | 0.6 | 2.9 | Y | Y | Y | N | N |
| R4 | Responder | Late post | 283 | 0 | 1.24 | 0.28 | <19 | >90 | 0.7 | 3.3 | Y | N | Y | N | N |
| R5 | Responder | Pretreatment | -108 | 10 | 0.59 | 0.1 | 79 | 60 | 1.7 | 3.6 | N | Y | Y | N | Y |
| R5 | Responder | Early post | 172 | 2 | 0.64 | 0.11 | 39 | 58 | 1.5 | 4.6 | Y | Y | Y | N | N |
| NR1 | Non-Responder | Pretreatment | -13 | 8 | 0.55 | 0.05 | 0 | >90 | 1.3 | 1.7 | N | N | Y | N | N |
| NR1 | Non-Responder | Early post | 127 | 0 | 0.72 | 0.12 | 6 | >90 | 1.4 | 1.7 | N | N | Y | N | N |
| NR1 | Non-Responder | Late post | 204 | 7 | 0.37 | 0.04 | 6 | >90 | 0.9 | 1.3 | N | N | Y | N | N |
| NR2 | Non-Responder | Pretreatment | 0 | 6 | 1.03 | 0.29 | 57 | >90 | 1.9 | 3.1 | N | N | N | N | N |
| NR2 | Non-Responder | Early post | 74 | 6 | 1.36 | 0.32 | 262 | >90 | 1.6 | 4.4 | N | N | N | N | N |
| NR2 | Non-Responder | Late post | 234 | 6 | 1.35 | 0.34 | 125 | >90 | 1.6 | 4.3 | N | Y | N | N | N |
| NR3 | Non-Responder | Pretreatment | -78 | 6 | 1.02 | 0.11 | 124 | 78 | 0.5 | 2.4 | N | N | N | N | N |
| NR3 | Non-Responder | Early post | 76 | 6 | 1.17 | 0.15 | 46 | 79 | 0.3 | 1.6 | Y | Y | N | N | N |
| NR4 | Non-Responder | Pretreatment | 0 | 4 | 0.86 | 0.28 | 60 | >90 | 0.7 | 1.7 | Y | Y | N | N | N |
| NR4 | Non-Responder | Early post | 124 | 4 | 0.95 | 0.32 | 131 | 89 | 0.7 | 1.7 | Y | N | N | N | N |

[1] Response to rituximab was defined by a clinician at 12 months post-treatment. Patients were classified as responders if clinical SLEDAI was ≤ 2 and required ≤ 4mg prednisolone once daily at 12 months. When the sole manifestation was proteinuria, patients were classified as responders when the patient was non-nephrotic with >50% reduction in urine protein : creatinine ratio (uPCR).

[2] clinical SLEDAI: modified SLE Disease Activity Index 2000 which does not take serology into account.

**Supplementary Table 3.** DEGs between pretreatment and late post-treatment naive B cells in responders. (FDR < 0.05, CPM = counts per million)

| Gene | $Log_2FC$ | Average logCPM | F | P-Value | FDR |
|---|---|---|---|---|---|
| *STAG3* | -2.7608609 | 7.16426389 | 121.34746 | 5.81E-26 | 8.01E-22 |
| *NETO1* | -4.6603395 | 5.07689482 | 91.4787762 | 1.17E-21 | 8.08E-18 |
| *MNDA* | -2.6179865 | 5.88408274 | 82.4583373 | 1.21E-19 | 5.57E-16 |
| *KCNG1* | -3.0421472 | 5.79928824 | 71.7998172 | 2.44E-17 | 8.40E-14 |
| *CD1C* | 2.05121118 | 5.91728394 | 58.7158768 | 1.85E-14 | 5.08E-11 |
| *CD9* | 2.20031171 | 5.76671538 | 58.364432 | 2.21E-14 | 5.08E-11 |
| *MIR4435-2HG* | -2.2998406 | 5.54092942 | 55.9005879 | 7.86E-14 | 1.55E-10 |
| *SLC38A11* | -2.0455174 | 6.11074096 | 54.3699291 | 1.68E-13 | 2.90E-10 |
| *THRB* | -2.9743706 | 5.28003083 | 52.8609458 | 4.54E-13 | 6.95E-10 |
| *JADE3* | -2.5631831 | 5.71685675 | 52.2271395 | 6.50E-13 | 8.96E-10 |
| *JCHAIN* | 1.33346695 | 8.31027876 | 47.5169657 | 5.54E-12 | 6.94E-09 |
| *SOX4* | 1.90110157 | 5.56577073 | 42.9215982 | 5.75E-11 | 6.26E-08 |
| *ELL2* | -1.6247155 | 6.41434383 | 42.8690846 | 5.91E-11 | 6.26E-08 |
| *RGS2* | -1.7734446 | 5.55725589 | 42.3090124 | 7.88E-11 | 7.75E-08 |
| *RAB31* | -1.8958664 | 5.19807281 | 40.0431598 | 2.50E-10 | 2.30E-07 |
| *SERPINB6* | -1.7264838 | 5.93032944 | 39.3612016 | 3.55E-10 | 2.95E-07 |
| *S100A8* | -1.7830561 | 5.54479369 | 39.3133827 | 3.64E-10 | 2.95E-07 |
| *LARGE1* | -1.5281701 | 6.58395037 | 37.8295568 | 7.77E-10 | 5.95E-07 |
| *AC007952.4* | -1.1826222 | 8.00948954 | 37.3747408 | 9.82E-10 | 6.95E-07 |
| *PPP1R14A* | 1.30600173 | 6.81050059 | 37.3007506 | 1.02E-09 | 6.95E-07 |
| *NR3C2* | -1.7614003 | 5.59948552 | 37.2243654 | 1.06E-09 | 6.95E-07 |
| *AKAP6* | -3.0431447 | 4.66348299 | 36.3729574 | 1.64E-09 | 1.03E-06 |
| *AC004687.1* | 1.24094354 | 7.19306657 | 35.7745686 | 2.23E-09 | 1.34E-06 |
| *AC233755.2* | -1.2629108 | 7.29855764 | 35.8382769 | 3.58E-09 | 1.97E-06 |
| *MYO1E* | 1.3622056 | 6.63859544 | 34.8582871 | 3.58E-09 | 1.97E-06 |
| *SESN1* | -1.2757616 | 7.26149532 | 33.6487084 | 6.64E-09 | 3.49E-06 |
| *NT5E* | -1.4653315 | 6.39465325 | 33.5927256 | 6.83E-09 | 3.49E-06 |
| *MPEG1* | 1.4970724 | 5.80777273 | 32.7985117 | 1.03E-08 | 5.06E-06 |
| *PRCD* | -2.0902018 | 4.94320975 | 31.5041152 | 2.00E-08 | 9.50E-06 |
| *PLD4* | 1.08836391 | 7.50808029 | 31.3480632 | 2.17E-08 | 9.95E-06 |
| *LYZ* | -1.4703774 | 5.77869315 | 30.8240963 | 2.84E-08 | 1.26E-05 |
| *PARM1* | -2.6854984 | 4.33690603 | 30.7506723 | 2.95E-08 | 1.27E-05 |
| *MIR181A1HG* | 1.77535045 | 5.19666129 | 30.6583188 | 3.09E-08 | 1.29E-05 |
| *CLEC4C* | 2.89037017 | 4.12640261 | 29.9850101 | 4.37E-08 | 1.77E-05 |
| *MTRNR2L8* | -1.5606775 | 5.80239761 | 32.9472794 | 4.57E-08 | 1.80E-05 |
| *TP53INP1* | -1.1448649 | 7.33827213 | 29.337806 | 7.24E-08 | 2.77E-05 |
| *CD83* | -1.0237824 | 8.92390674 | 28.8653331 | 7.99E-08 | 2.98E-05 |
| *UGT8* | -2.0000111 | 5.08011591 | 28.544596 | 9.19E-08 | 3.33E-05 |
| *CCDC191* | 1.18182196 | 6.8800179 | 28.460995 | 9.60E-08 | 3.39E-05 |
| *ZFP36L2* | -1.129022 | 9.95189399 | 28.3693067 | 1.03E-07 | 3.55E-05 |
| *PRDM1* | -2.3727186 | 4.3780293 | 27.7304531 | 1.40E-07 | 4.70E-05 |
| *LINC01374* | 1.6762121 | 5.0022223 | 27.5580083 | 1.54E-07 | 5.05E-05 |
| *THEMIS2* | -1.8452483 | 5.31256492 | 28.1049446 | 2.24E-07 | 7.18E-05 |
| *PTGER4* | -1.259481 | 6.24670144 | 26.4276834 | 2.75E-07 | 8.60E-05 |
| *IKZF2* | 1.36794836 | 5.6010455 | 25.9910302 | 3.44E-07 | 0.00010536 |
| *BCL2A1* | -1.2182109 | 6.12129203 | 25.4242154 | 4.62E-07 | 0.00013824 |
| *AC002460.2* | -3.7543614 | 3.67234775 | 25.1744679 | 5.25E-07 | 0.00015399 |
| *SELENOM* | -1.4859975 | 5.36499659 | 24.7806412 | 6.44E-07 | 0.00018494 |
| *CEMIP2* | -0.9384255 | 7.78703806 | 24.4316841 | 7.72E-07 | 0.00021711 |
| *TUBB6* | 1.24713082 | 5.87340392 | 24.2304985 | 8.57E-07 | 0.00023618 |
| *AIM2* | -3.2698161 | 3.89928048 | 23.8755311 | 1.03E-06 | 0.0002784 |
| *KLF9* | -1.1552684 | 6.32272224 | 23.5168268 | 1.24E-06 | 0.00032985 |
| *AC108879.1* | 1.27464829 | 6.17206203 | 23.3149043 | 1.38E-06 | 0.00035846 |
| *SYNE2* | 0.92041166 | 7.5285719 | 22.8317326 | 1.77E-06 | 0.00045229 |
| *ZNF208* | -2.3727404 | 4.10381104 | 22.204739 | 2.46E-06 | 0.00061537 |

| | | | | | |
|---|---|---|---|---|---|
| *TTN* | 0.9471973 | 7.05570132 | 21.9531335 | 2.80E-06 | 0.00068899 |
| *AC245060.5* | 1.22762004 | 5.7933848 | 21.7513202 | 3.11E-06 | 0.00075194 |
| *PTPRJ* | 1.04595323 | 6.35173114 | 21.5624047 | 3.43E-06 | 0.00081543 |
| *JUNB* | -1.0026651 | 10.7282438 | 21.5846011 | 3.85E-06 | 0.0008986 |
| *ZHX3* | -1.1093114 | 6.04082597 | 21.1977414 | 4.16E-06 | 0.00095417 |
| *PLAG1* | -3.3153563 | 3.7034437 | 20.9013406 | 4.85E-06 | 0.00109449 |
| *KCNQ5* | 1.00320261 | 6.68907391 | 20.7994323 | 5.11E-06 | 0.00113566 |
| *LSR* | -1.5743939 | 5.0841009 | 20.7020124 | 5.38E-06 | 0.00116657 |
| *AL139020.1* | 0.89948866 | 7.36379548 | 20.6871748 | 5.42E-06 | 0.00116657 |
| *WASF1* | 1.5855574 | 4.8523104 | 20.4167373 | 6.28E-06 | 0.00131333 |
| *HMCES* | 0.98223639 | 6.78348216 | 20.4013316 | 6.29E-06 | 0.00131333 |
| *UACA* | -1.8011055 | 5.11310422 | 21.2835821 | 6.50E-06 | 0.00133721 |
| *PRDM4* | -1.017732 | 6.33476881 | 20.2634931 | 6.76E-06 | 0.00136988 |
| *AEBP1* | 1.45681647 | 4.98323545 | 20.0676389 | 7.49E-06 | 0.00149552 |
| *GLUL* | 1.22185216 | 5.56902381 | 20.0363077 | 7.61E-06 | 0.0014985 |
| *FAM81A* | 1.73563038 | 4.46005887 | 19.9697533 | 7.88E-06 | 0.00151858 |
| *BTLA* | -0.847435 | 7.91568221 | 19.9569613 | 7.94E-06 | 0.00151858 |
| *Z93241.1* | -1.2783231 | 5.21172693 | 19.9252485 | 8.07E-06 | 0.00152282 |
| *LMO2* | -1.2743762 | 5.53186938 | 19.7418238 | 8.88E-06 | 0.00165347 |
| *CNTNAP2* | -1.1397668 | 6.08730451 | 19.4455074 | 1.04E-05 | 0.00190502 |
| *ST6GALNAC3* | -1.0171545 | 6.7708781 | 19.2029341 | 1.18E-05 | 0.00213452 |
| *MLLT3* | -1.4470346 | 5.08103105 | 18.7493328 | 1.49E-05 | 0.00267199 |
| *GNA12* | -0.9654787 | 6.67560216 | 18.683519 | 1.55E-05 | 0.00273033 |
| *AL079338.1* | 1.29545947 | 5.16543745 | 18.6414807 | 1.58E-05 | 0.00275585 |
| *CD69* | -0.9206435 | 10.7229954 | 18.6480117 | 1.63E-05 | 0.00277876 |
| *ZNF669* | -1.2700553 | 5.25837309 | 18.5780112 | 1.63E-05 | 0.00277876 |
| *PRKN* | -1.5306563 | 4.98879143 | 18.3159286 | 1.87E-05 | 0.00312498 |
| *CSGALNACT1* | 0.88709596 | 6.67185051 | 18.3076707 | 1.88E-05 | 0.00312498 |
| *HSPH1* | -0.979611 | 6.53978821 | 18.2492776 | 1.94E-05 | 0.00315697 |
| *DDIT4* | -1.2329437 | 6.11224975 | 18.8374835 | 1.95E-05 | 0.00315697 |
| *FRY* | 1.67853271 | 4.48614407 | 18.2002618 | 1.99E-05 | 0.00319086 |
| *BACE2* | -1.2867455 | 5.64577558 | 18.1365935 | 2.06E-05 | 0.00326138 |
| *MS4A7* | 1.12765645 | 5.4419924 | 17.8456995 | 2.40E-05 | 0.00375649 |
| *SOCS3* | -2.0078501 | 4.09177372 | 17.6839697 | 2.61E-05 | 0.00404373 |
| *SNHG5* | -0.7792632 | 8.1893143 | 17.466524 | 2.93E-05 | 0.00448307 |
| *PPBP* | 1.14064951 | 5.80204921 | 17.1902868 | 3.39E-05 | 0.00507914 |
| *BCL2L11* | -1.2087386 | 5.68268055 | 17.1874371 | 3.39E-05 | 0.00507914 |
| *TTC28* | 1.45736776 | 4.65032349 | 17.0300115 | 3.68E-05 | 0.0054586 |
| *HCK* | 0.98268567 | 5.95623192 | 16.8641321 | 4.03E-05 | 0.00590133 |
| *IRAK3* | -1.6421351 | 4.77234795 | 16.6255518 | 4.56E-05 | 0.00661257 |
| *BRCA2* | -1.473 | 5.06385967 | 16.5914495 | 4.64E-05 | 0.00666237 |
| *CD3D* | -2.2617849 | 4.07175086 | 16.2723398 | 5.49E-05 | 0.00776783 |
| *AC104971.3* | 0.92654285 | 6.4935673 | 16.2612575 | 5.53E-05 | 0.00776783 |
| *PIK3IP1* | -0.8262503 | 7.28709629 | 16.2028182 | 5.70E-05 | 0.00793018 |
| *RASL11A* | -3.1937841 | 4.04038961 | 16.1741192 | 5.79E-05 | 0.00797069 |
| *AC020659.1* | -2.6238007 | 3.69577317 | 16.0911396 | 6.04E-05 | 0.00824511 |
| *ZNF331* | -0.8276605 | 7.02563778 | 15.9168138 | 6.63E-05 | 0.00895148 |
| *GNG11* | 1.57527642 | 4.50846691 | 15.8853928 | 6.74E-05 | 0.00901293 |
| *CCDC112* | 0.98044583 | 6.08049035 | 15.8636664 | 6.82E-05 | 0.0090293 |
| *AC005165.1* | -2.7956577 | 3.53016514 | 15.7236251 | 7.34E-05 | 0.00963017 |
| *COL9A3* | 1.38270692 | 4.87880406 | 15.5777136 | 7.93E-05 | 0.01030423 |
| *EML6* | 2.91470097 | 3.50119583 | 15.4435918 | 8.51E-05 | 0.0109583 |
| *FGD4* | 2.41675269 | 3.71435285 | 15.0392878 | 0.00010542 | 0.01344767 |
| *PTPRN2* | -1.1601854 | 5.34153641 | 14.9565404 | 0.00011014 | 0.01392139 |
| *WDR49* | 1.93751665 | 3.94257921 | 14.820343 | 0.00011839 | 0.01482736 |
| *TYROBP* | -1.3810472 | 4.80818714 | 14.7926599 | 0.00012014 | 0.01491101 |
| *ADGRG5* | 1.50082599 | 4.57698747 | 14.5734907 | 0.00013495 | 0.01659944 |
| *SYTL3* | -2.1992773 | 4.00078446 | 14.4906598 | 0.00014101 | 0.01719181 |
| *ITGB2-AS1* | 0.97697891 | 5.96324136 | 14.4728651 | 0.00014235 | 0.01720271 |
| *FCRL5* | -0.8096103 | 7.19351486 | 14.4099689 | 0.00014728 | 0.01760371 |
| *ANKRD37* | -1.2748768 | 4.84996399 | 14.3966932 | 0.00014822 | 0.01760371 |

| | | | | | |
|---|---|---|---|---|---|
| *HERPUD1* | -0.771295 | 7.91868049 | 14.6886395 | 0.00015448 | 0.01804622 |
| *CCR7* | -0.7210947 | 8.3665909 | 14.3090339 | 0.00015528 | 0.01804622 |
| *AUTS2* | 0.70607641 | 7.87488475 | 14.3018545 | 0.00015588 | 0.01804622 |
| *CCND3* | -0.741042 | 8.83810036 | 14.3002527 | 0.00015889 | 0.0181981 |
| *RESF1* | -0.7151338 | 8.67442897 | 14.2546953 | 0.00015983 | 0.0181981 |
| *CXCR4* | -0.8021704 | 10.8990049 | 14.1784546 | 0.00016644 | 0.018795 |
| *POU6F1* | -4.5075645 | 3.30591749 | 14.1608076 | 0.000168 | 0.01881782 |
| *CCNG2* | 0.81530118 | 6.82865775 | 14.0789224 | 0.00017548 | 0.01949632 |
| *GPR183* | -0.7791739 | 6.81224106 | 14.0241313 | 0.00018066 | 0.01983266 |
| *KLF2* | -0.8257834 | 11.3603795 | 14.0166503 | 0.00018138 | 0.01983266 |
| *CHPT1* | -0.7136958 | 8.33464266 | 14.0010706 | 0.00018289 | 0.01984016 |
| *LZTFL1* | 1.12934364 | 5.16321054 | 13.9336633 | 0.00018957 | 0.02040358 |
| *SUGCT* | 2.09860646 | 4.36740223 | 13.8881783 | 0.00019421 | 0.02074119 |
| *AEN* | -1.1509056 | 5.1868394 | 13.7692737 | 0.00020689 | 0.02192581 |
| *ACTG1* | 0.7696148 | 10.1028866 | 13.6351694 | 0.0002222 | 0.02336852 |
| *PEG10* | -1.3403164 | 4.68662168 | 13.5311743 | 0.00023486 | 0.02451224 |
| *RHOC* | 0.97283103 | 5.70798258 | 13.4416594 | 0.00024633 | 0.02551635 |
| *ARID5A* | -0.9356031 | 5.98392922 | 13.3934793 | 0.00025274 | 0.02598467 |
| *SYN3* | -4.5449824 | 3.36813407 | 13.3689634 | 0.00025606 | 0.02602466 |
| *INSIG2* | -0.9620398 | 5.82654443 | 13.3628009 | 0.0002569 | 0.02602466 |
| *AIF1* | -1.2904634 | 4.75067676 | 13.1367765 | 0.00028982 | 0.0291445 |
| *CYCS* | -0.679119 | 7.82793964 | 13.0119677 | 0.00030983 | 0.030931 |
| *MRPL50* | -1.0340899 | 5.29748833 | 12.992636 | 0.00031299 | 0.03102243 |
| *ADAM28* | -0.697522 | 8.87461183 | 12.9622056 | 0.00031812 | 0.03130541 |
| *TNS3* | 1.18177001 | 4.9121998 | 12.8444126 | 0.00033878 | 0.03310197 |
| *PLIN2* | -1.2562838 | 4.81936524 | 12.7835835 | 0.00034997 | 0.03390614 |
| *ZNF318* | -0.6930062 | 7.69422365 | 12.7702509 | 0.00035273 | 0.03390614 |
| *CYTH3* | 1.90404949 | 3.89141901 | 12.7573285 | 0.00035492 | 0.03390614 |
| *CCDC200* | -0.915653 | 6.24600896 | 12.7489518 | 0.00035686 | 0.03390614 |
| *LINC02161* | -1.7489386 | 5.11459344 | 12.7138569 | 0.00036326 | 0.03426953 |
| *SOCS1* | -0.9050601 | 5.93912196 | 12.7015846 | 0.00036565 | 0.03426953 |
| *VCAN* | -1.8122066 | 4.04877268 | 12.6717926 | 0.00037153 | 0.03458452 |
| *IQGAP2* | 0.86046489 | 5.99695305 | 12.5493826 | 0.0003968 | 0.0365239 |
| *SMAP2* | -0.7219052 | 9.6040227 | 12.5446901 | 0.00039766 | 0.0365239 |
| *GRASP* | -0.9647408 | 5.72208545 | 12.4861577 | 0.00041031 | 0.03743639 |
| *CFLAR* | -0.7202961 | 7.06666598 | 12.4052447 | 0.00042848 | 0.03883625 |
| *ANKUB1* | 0.91455542 | 5.69255215 | 12.3806122 | 0.00043416 | 0.03909464 |
| *ARRDC2* | -0.7986509 | 6.48684666 | 12.347381 | 0.00044196 | 0.03953814 |
| *ARHGAP32* | 0.97273753 | 5.5776374 | 12.3336359 | 0.00044523 | 0.0395733 |
| *JUN* | -0.7373838 | 10.7415451 | 12.2576975 | 0.00046376 | 0.0408648 |
| *SLC50A1* | 0.71299761 | 7.15094971 | 12.2497521 | 0.00046569 | 0.0408648 |
| *AMDHD2* | 1.30542369 | 4.93460463 | 12.162159 | 0.00048807 | 0.04255774 |
| *PLCL2* | 0.6493396 | 8.18312077 | 11.98854 | 0.00053569 | 0.04627188 |
| *CLNS1A* | -0.671226 | 7.51749798 | 11.9826752 | 0.00053738 | 0.04627188 |
| *SPRY1* | -1.5992071 | 4.22802986 | 11.9215731 | 0.00055529 | 0.04746271 |
| *MGAT5* | -0.6471746 | 8.2425563 | 11.9121726 | 0.0005581 | 0.04746271 |
| *XBP1* | -0.9072989 | 5.69061031 | 11.8768887 | 0.00056877 | 0.04758788 |
| *LHFPL2* | 1.14426822 | 4.93587275 | 11.8730257 | 0.00056995 | 0.04758788 |
| *AC012447.1* | -1.334816 | 4.27945896 | 11.862473 | 0.00057319 | 0.04758788 |
| *GDF11* | 1.39068785 | 4.2438082 | 11.85473 | 0.00057558 | 0.04758788 |
| *HLA-DRB1* | -0.7760297 | 11.7998603 | 11.847577 | 0.00057779 | 0.04758788 |
| *SLC35F1* | 1.08298444 | 5.07703935 | 11.8358877 | 0.00058143 | 0.04758788 |
| *C2orf49* | -0.9879718 | 5.61156847 | 11.8228127 | 0.00058553 | 0.04758788 |
| *PPP1R13B* | -2.5987791 | 3.78188721 | 11.8174776 | 0.00058721 | 0.04758788 |
| *RNASE6* | -0.6504752 | 7.85356037 | 11.7633842 | 0.00060477 | 0.04872457 |

**Supplementary Table 4.** DEGs between pretreatment and early post-treatment non-B cells that change differently between responders and non-responders. (CPM = counts per million)

| Cell type | Gene | $Log_2FC$ | logCPM | F | FDR | Responder effect size | Non-responder effect size |
|---|---|---|---|---|---|---|---|
| Naive CD4 T | *SCGB3A1* | -2.2770 | 6.1099 | 86.6178 | 1.520E-08 | -1.0249 | 0.5534 |
| Naive CD4 T | *ZFP36* | 0.7867 | 7.9532 | 32.5896 | 1.362E-02 | 0.2783 | -0.2670 |
| Naive CD4 T | *TAGAP* | -0.6704 | 8.6987 | 28.5061 | 3.000E-02 | -0.2216 | 0.2430 |
| CD4 TCM | *GZMK* | 4.9273 | 6.0972 | 137.4109 | 1.520E-27 | 1.6439 | -1.7714 |
| CD4 TCM | *FOS* | 1.5130 | 8.4480 | 73.5956 | 9.560E-13 | 0.7096 | -0.3392 |
| CD4 TCM | *TENT5C* | 1.9331 | 6.6007 | 62.0691 | 1.600E-11 | 0.6134 | -0.7265 |
| CD4 TCM | *ZFP36* | 1.0882 | 8.5229 | 43.2570 | 1.990E-07 | 0.2733 | -0.4810 |
| CD4 TCM | *FAM13A* | -1.2254 | 7.1634 | 31.1298 | 7.060E-05 | -0.5920 | 0.2573 |
| CD4 TCM | *CYTOR* | 1.4855 | 6.3308 | 28.9333 | 1.671E-04 | 0.4563 | -0.5734 |
| CD4 TCM | *BTG2* | 0.9712 | 8.0799 | 28.7718 | 1.671E-04 | 0.4521 | -0.2210 |
| CD4 TCM | *GADD45B* | 1.2694 | 6.8628 | 28.1422 | 1.932E-04 | 0.8773 | -0.0026 |
| CD4 TCM | *NR4A2* | 1.6375 | 5.9405 | 28.0027 | 1.932E-04 | 0.9004 | -0.2346 |
| CD4 TCM | *IER2* | 1.0812 | 8.1454 | 29.6958 | 3.966E-04 | 0.6350 | -0.1145 |
| CD4 TCM | *TRIM22* | -0.7817 | 8.8326 | 24.0701 | 1.208E-03 | -0.4897 | 0.0521 |
| CD4 TCM | *SLC2A3* | 0.8139 | 8.5011 | 22.9547 | 1.977E-03 | 0.3262 | -0.2380 |
| CD4 TCM | *IRF1* | 0.8794 | 7.9720 | 22.5789 | 2.219E-03 | 0.6981 | 0.0885 |
| CD4 TCM | *PTGER2* | -1.0047 | 7.2119 | 22.3635 | 2.271E-03 | -0.5947 | 0.1017 |
| CD4 TCM | *IL2RA* | -1.9568 | 5.3453 | 22.2638 | 2.271E-03 | -1.1716 | 0.1847 |
| CD4 TCM | *ANXA1* | 0.7319 | 8.9526 | 21.3719 | 3.463E-03 | 0.4022 | -0.1051 |
| CD4 TCM | *SOCS2* | -1.6784 | 5.6841 | 21.2033 | 3.476E-03 | -0.9068 | 0.2566 |
| CD4 TCM | *IFNGR2* | -1.4126 | 6.0448 | 19.9776 | 6.247E-03 | -0.7348 | 0.2443 |
| CD4 TCM | *MIR4435-2HG* | 1.6707 | 5.5347 | 19.5804 | 7.258E-03 | 0.7279 | -0.4302 |
| CD4 TCM | *DNAJB1* | 0.7739 | 8.1293 | 19.3135 | 7.933E-03 | 0.4283 | -0.1082 |
| CD4 TCM | *OSM* | 2.7046 | 4.4055 | 18.3660 | 1.241E-02 | 1.4952 | -0.3795 |
| CD4 TCM | *SMAD7* | 7.1461 | 3.3578 | 18.2522 | 1.257E-02 | 4.2604 | -0.6929 |
| CD4 TCM | *JUND* | 0.6975 | 9.8317 | 18.7845 | 2.123E-02 | 0.3499 | -0.1335 |
| CD4 TCM | *PLAC8* | -0.7420 | 8.0367 | 17.0901 | 2.123E-02 | -0.2058 | 0.3085 |
| CD4 TCM | *ICOS* | 0.7115 | 8.2952 | 16.4372 | 2.948E-02 | 0.2946 | -0.1986 |
| CD4 TCM | *PUDP* | -1.5181 | 5.4719 | 16.2771 | 3.008E-02 | -0.7420 | 0.3103 |
| CD4 TCM | *FBXO33* | 0.9925 | 6.5628 | 15.7962 | 3.734E-02 | 0.4864 | -0.2015 |
| CD4 TCM | *PCLAF* | 4.6491 | 3.1556 | 15.6993 | 3.792E-02 | 2.7527 | -0.4698 |
| CD4 TCM | *VAV3* | -1.6597 | 5.1914 | 15.7148 | 3.841E-02 | -1.0424 | 0.1081 |
| CD4 TEM | *GADD45B* | 1.0816 | 7.3047 | 33.6102 | 3.639E-03 | 0.6308 | -0.1189 |
| CD4 TEM | *SELL* | -0.7872 | 9.2141 | 26.7606 | 1.791E-02 | -0.3737 | 0.1719 |
| CD4 TEM | *SCGB3A1* | -2.3991 | 4.2707 | 23.1126 | 4.432E-02 | -0.9816 | 0.6813 |
| Treg | *S100A8* | 2.7025 | 5.5833 | 51.8084 | 1.050E-08 | 1.0900 | -0.7833 |
| Treg | *FOS* | 1.1311 | 7.1874 | 25.4625 | 3.182E-03 | 0.4712 | -0.3128 |
| Treg | *BTG2* | 0.9723 | 7.8194 | 23.1862 | 7.602E-03 | 0.5441 | -0.1298 |
| Treg | *ERAP2* | -1.2285 | 6.6983 | 22.4486 | 7.602E-03 | -0.7741 | 0.0774 |
| Treg | *RNF19A* | -0.8687 | 7.8742 | 20.7967 | 1.317E-02 | -0.6802 | -0.0781 |
| Treg | *MCL1* | 0.8513 | 8.1001 | 20.1809 | 1.317E-02 | 0.3493 | -0.2407 |
| Treg | *CST3* | 1.9231 | 5.0145 | 20.1294 | 1.317E-02 | 0.8992 | -0.4338 |
| Treg | *LMNA* | 1.5855 | 5.6107 | 20.0669 | 1.317E-02 | 0.8612 | -0.2377 |
| Treg | *IL6ST* | -0.8480 | 7.9580 | 19.4864 | 1.586E-02 | -0.5782 | 0.0096 |
| Treg | *IER2* | 0.9590 | 8.2178 | 20.0574 | 3.089E-02 | 0.5601 | -0.1046 |
| Treg | *TENT5C* | 0.9565 | 6.9741 | 16.9661 | 4.871E-02 | 0.2612 | -0.4018 |
| Naive CD8 T | *SCGB3A1* | -1.4140 | 6.6110 | 46.3777 | 6.820E-05 | -0.6855 | 0.2946 |
| Naive CD8 T | *TAGAP* | -0.6405 | 8.3393 | 27.3993 | 1.291E-02 | -0.3076 | 0.1364 |
| Naive CD8 T | *GADD45B* | 0.7706 | 6.8759 | 24.1478 | 2.901E-02 | 0.4892 | -0.0450 |
| Naive CD8 T | *BTG2* | 0.9432 | 7.3720 | 29.8325 | 3.861E-02 | 0.4614 | -0.1924 |
| Naive CD8 T | *TRIM22* | -0.5732 | 8.5759 | 22.1292 | 4.245E-02 | -0.4746 | -0.0773 |
| Naive CD8 T | *ZFP36* | 0.8150 | 8.0469 | 27.3967 | 4.347E-02 | 0.2989 | -0.2660 |
| CD8 TEM | *SOCS3* | 1.6832 | 6.9077 | 34.4294 | 6.480E-05 | 0.8993 | -0.2674 |
| CD8 TEM | *TENT5C* | 1.4699 | 6.9550 | 29.2612 | 4.542E-04 | 0.5206 | -0.4983 |

| | | | | | | | |
|---|---|---|---|---|---|---|---|
| CD8 TEM | *TXNIP* | -0.8205 | 11.3339 | 27.1203 | 9.144E-04 | -0.5741 | -0.0053 |
| CD8 TEM | *MYB* | -5.7458 | 4.1138 | 25.5911 | 1.512E-03 | -3.6266 | 0.3560 |
| CD8 TEM | *IER2* | 0.9984 | 8.4498 | 22.3095 | 6.650E-03 | 0.5397 | -0.1523 |
| CD8 TEM | *TOB1* | -1.1484 | 7.2379 | 21.1353 | 1.050E-02 | -0.7855 | 0.0105 |
| TEMRA | *NR4A2* | 1.5171 | 6.7647 | 47.6292 | 3.070E-08 | 0.4459 | -0.6057 |
| TEMRA | *LGALS1* | -0.6915 | 9.5554 | 28.7065 | 3.015E-04 | -0.4938 | -0.0145 |
| TEMRA | *CD69* | 0.7479 | 8.8299 | 27.8889 | 3.564E-04 | 0.1668 | -0.3516 |
| TEMRA | *ZNF331* | 1.1751 | 6.3396 | 22.2495 | 4.410E-03 | 0.5621 | -0.2524 |
| TEMRA | *S100A9* | 1.6353 | 5.5337 | 22.9165 | 4.410E-03 | 0.3600 | -0.7735 |
| TEMRA | *PRF1* | -0.5548 | 10.2861 | 21.9153 | 4.410E-03 | -0.2794 | 0.1051 |
| TEMRA | *GNLY* | -0.4745 | 12.6926 | 21.9128 | 4.410E-03 | -0.2668 | 0.0621 |
| TEMRA | *BTG2* | 0.9015 | 7.0432 | 21.2510 | 5.608E-03 | 0.2819 | -0.3430 |
| TEMRA | *JUND* | 0.5728 | 9.5708 | 20.4365 | 7.799E-03 | 0.2396 | -0.1575 |
| TEMRA | *RHOB* | 1.1135 | 6.1463 | 19.4814 | 1.178E-02 | 0.6604 | -0.1114 |
| TEMRA | *KLRC4* | -0.8831 | 7.1077 | 19.4315 | 1.178E-02 | -0.1979 | 0.4142 |
| TEMRA | *TENT5C* | 0.8069 | 7.0394 | 16.6637 | 4.413E-02 | 0.1412 | -0.4181 |
| DN T | *LGALS1* | 1.9243 | 8.7170 | 49.9140 | 3.090E-08 | 0.3405 | -0.9933 |
| DN T | *PRF1* | 2.6981 | 7.2727 | 46.2985 | 6.480E-08 | 1.3048 | -0.5653 |
| DN T | *IL7R* | -1.4253 | 9.4760 | 34.9179 | 1.340E-05 | -0.4719 | 0.5161 |
| DN T | *CD74* | 1.2530 | 10.6437 | 34.8994 | 1.340E-05 | 0.3713 | -0.4972 |
| DN T | *GZMA* | 1.4240 | 9.3509 | 30.0551 | 1.067E-04 | 0.5984 | -0.3886 |
| DN T | *HLA-DRA* | 1.9073 | 7.2959 | 28.4851 | 2.503E-04 | 0.4549 | -0.8671 |
| DN T | *HLA-DPB1* | 1.7203 | 7.7195 | 26.7100 | 3.757E-04 | 0.6045 | -0.5879 |
| DN T | *ANXA5* | 1.6618 | 7.5568 | 25.7579 | 5.459E-04 | 0.6671 | -0.4848 |
| DN T | *S100A4* | 1.0202 | 10.4810 | 23.1344 | 1.917E-03 | 0.1734 | -0.5338 |
| DN T | *CD70* | 6.2819 | 4.9018 | 22.8181 | 2.054E-03 | 2.7073 | -1.6470 |
| DN T | *CCL4* | 9.6156 | 4.3962 | 22.1337 | 2.688E-03 | 5.5176 | -1.1474 |
| DN T | *GAPDH* | 0.8731 | 10.9724 | 18.4944 | 1.665E-02 | 0.1828 | -0.4224 |
| DN T | *COTL1* | 1.0015 | 9.4926 | 17.5757 | 2.586E-02 | 0.2259 | -0.4683 |
| DN T | *HMGB2* | 1.1796 | 8.1932 | 16.9015 | 3.752E-02 | 0.4699 | -0.3477 |
| DN T | *HLA-DPA1* | 1.6007 | 7.1732 | 16.0496 | 4.888E-02 | 0.6135 | -0.4960 |
| CD56Dim NK | *LGALS3* | -1.4737 | 7.2694 | 46.3252 | 1.380E-07 | -0.7435 | 0.2780 |
| CD56Dim NK | *HES4* | -2.1797 | 5.8554 | 29.0730 | 4.768E-04 | -1.5317 | -0.0209 |
| CD56Dim NK | *AC243829.2* | -1.7025 | 6.2819 | 27.1441 | 1.667E-03 | -1.0273 | 0.1528 |
| CD56Dim NK | *S100A8* | 1.9561 | 5.2761 | 23.8272 | 3.104E-03 | 0.5329 | -0.8230 |
| CD56Dim NK | *IL32* | -0.7252 | 11.2532 | 23.8909 | 3.104E-03 | -0.3538 | 0.1488 |
| CD56Dim NK | *S100A9* | 1.8214 | 5.2596 | 23.3440 | 3.104E-03 | 0.4163 | -0.8462 |
| CD56Dim NK | *LGALS1* | -0.9146 | 10.3643 | 24.3001 | 1.014E-02 | -0.6410 | -0.0070 |
| CD56Dim NK | *TRG-AS1* | -1.1504 | 6.8906 | 20.4277 | 1.014E-02 | -0.4334 | 0.3640 |
| CD56Dim NK | *UTS2* | 2.3033 | 5.1794 | 20.2807 | 1.014E-02 | 0.9393 | -0.6573 |
| CD56Dim NK | *XCL2* | 1.1210 | 7.2297 | 18.8796 | 2.735E-02 | 0.9476 | 0.1705 |
| CD56Dim NK | *IER2* | 0.8804 | 8.9301 | 19.8313 | 2.965E-02 | 0.4166 | -0.1936 |
| CD56Dim NK | *SOCS1* | -2.1123 | 4.7991 | 17.5500 | 2.965E-02 | -1.3858 | 0.0783 |
| CD56Dim NK | *LAG3* | -1.8449 | 5.2992 | 17.7991 | 2.965E-02 | -1.2122 | 0.0666 |
| CD56Dim NK | *TSC22D3* | -1.0121 | 8.3443 | 19.6579 | 3.193E-02 | -0.7029 | -0.0013 |
